# A graphical pipeline platform for MRS data processing and analysis: MRspecLAB

**DOI:** 10.1101/2025.04.30.25326661

**Authors:** Ying Xiao, Antonia Kaiser, Matthias Kockisch, Alex Back, Robin Carlet, Xinyu Liu, Zhiwei Huang, André Döring, Mark Widmaier, Lijing Xin

## Abstract

Magnetic resonance spectroscopy (MRS) and magnetic resonance spectroscopic imaging (MRSI), are non-invasive techniques used to quantify biochemical compounds in tissue, such as choline, creatine, glutamate, glutamine, γ-aminobutyric acid, N-acetylaspartate, etc. However, reliable quantification of MRS and MRSI data is challenging due to the complex processing steps involved, often requiring advanced expertise. Existing data processing software solutions often demand MRS expertise or coding knowledge, presenting a steep learning curve for novel users. Mastering these tools typically requires a long training time, which can be a barrier for users with limited technical backgrounds.

To address these challenges and create a tool that serves researchers using MRS/MRSI with a broad range of backgrounds, we developed MRspecLAB—an open-access, user-friendly software platform for MRS and MRSI data analysis. MRspecLAB is designed for easy installation and features an intuitive graphical pipeline editor that supports both predefined and customizable workflows. It also serves as a platform offering standardized pipelines while allowing users to integrate in-house functions for additional flexibility. Importantly, MRspecLAB is envisioned as an open platform beyond the MRS community, bridging the gap between technical experts and practitioners. It facilitates contributions, collaboration, and the sharing of data workflows and processing methodologies for diverse MRS/MRSI applications, supporting reproducibility practices.

## 1 Introduction

Magnetic resonance spectroscopy (MRS) and magnetic resonance spectroscopic imaging (MRSI), are non-invasive techniques for quantifying biochemical compounds in tissues, widely used to study neurochemicals such as choline, creatine, glutamate, glutamine, γ-aminobutyric acid, N-acetylaspartate, etc^1,2^. ^1^H MRS and MRSI are commonly applied^3^, and advances in hardware and acquisition methods have expanded MRS applications beyond ^1^H to include other nuclei such as deuterium (^2^H)^4^, carbon (^13^C)^5-7^, and phosphorus (^31^P)^8,9^. These innovations enable the exploration of molecular dynamics and have increased the applicability of MRS/MRSI in studying brain disorders^10^, including epilepsy^11^, stroke^12^, Alzheimer’s disease^13,14^, and tumors^15,16^.

However, accurate quantification of MRS and MRSI data is not straightforward as multiple spectral preprocessing steps are required to generate high-quality, analyzable spectra^17^. These steps generally include coil combination, frequency and phase correction, eddy current correction, removal of outlier spectra, apodization, and zero filling, etc.^18^ Each of these steps are essential for optimizing the SNR, minimizing artifacts and contaminations, and ensuring reliable and reproducible quantifications. Moreover, the preprocessing workflow is not uniform; different datasets, experimental setups, and applications may require specific adjustments and tailored preprocessing steps to address their unique characteristics. This variability adds to the technical complexity, making high-quality spectral processing a non-trivial task, and often requiring advanced expertise, specialized training, and in-depth knowledge of MRS principles.

Several well-established tools in the MRS community, such as FID-A^19^, FSL-MRS^20^, INSPECTOR^21^, Tarquin^22^, Osprey^23^, and more, provide robust support for MRS data simulation, processing, and quantification. These tools have significantly advanced the MRS field by offering reliable workflows and complete algorithms for spectral analysis. However, many of them are designed for in-depth analysis for users with significant knowledge about MRS data processing. For instance, the lack of intuitive graphical user interfaces (GUI) in some tools makes them less approachable, as they often rely on command-line operations or coding. Especially, a GUI-based pipeline editor is currently lacking, which would enable users to create processing workflows without requiring extensive programming skills. Additionally, some are MATLAB-based, requiring paid licenses, which may not be accessible to all users. Moreover, the setup and configuration process for some of the software or toolboxes can be time-consuming and may require users to have in-depth knowledge of their computer systems. Furthermore, most tools are designed for processing ^1^H MRS data, and provide limited support for X-nuclei data. As MRS expands to include a broader range of nuclei and applications, the demand for software capable of handling these data types efficiently has grown, highlighting the need for more versatile, accessible, and collaborative solutions, especially for clinical applications.

To address these needs and facilitate the application of advanced MRS and MRSI techniques for clinical neuroscientists, we developed MRspecLAB, an open-access, user-friendly, and collaborative software platform that can accommodate diverse datasets, minimize technical barriers for the wider community beyond the current MRS specialists, and facilitate the seamless sharing of functionalities and workflows in a straight-forward, plug and play manner. MRspecLAB offers:

1. **User-friendly GUI design**: An intuitive graphical user interface, including a graphic pipeline editor, enables drag-and-drop workflow creation.
2. **Platform character**: The modular coding structure enables the straightforward development of new data processing nodes with minimal programming expertise. The customer nodes can be easily integrated into the pipeline and shared with the community.
3. **Efficiency**: Batch processing supports rapid, reproducible analysis of large datasets, ideal for cohort studies.
4. **Open source**: Free to use, with available source code to encourage community contributions.
5. **Comprehensive functionality**: Supports various input formats from different vendors, different sequences, X-nuclei, and MRSI.

## 2 Methods

MRspecLAB was developed using Python 3 (RRID:SCR_008394), using free toolkits and libraries such as wxPython^24^, matplotlib^25^, pandas^26^, gswidgetkit^27^, gsnodegraph^28^, suspect^29^, pyMapVBVD^30^, etc. LCModel^31^ (RRID:SCR_014455), a widely recognized gold standard linear combination method in the MRS field for accurate metabolite quantification^18^, was integrated as the default fitting method to quantify metabolites. MRspecLAB is available as an easy-to-install precompiled version for windows, or as source-code package runnable on any Python prepared environment (https://github.com/MRSEPFL/MRspecLAB). More information about how to use the platform can be found in the user manual on github: https://github.com/MRSEPFL/MRspecLAB/blob/main/MANUAL.md.

### 2.1 User interface

MRspecLAB features an intuitive graphic interface designed to largely simplify the data analysis procedures for users of all expertise levels. Figure 1 provides an overview of the main window of the software:

**Figure 1:**
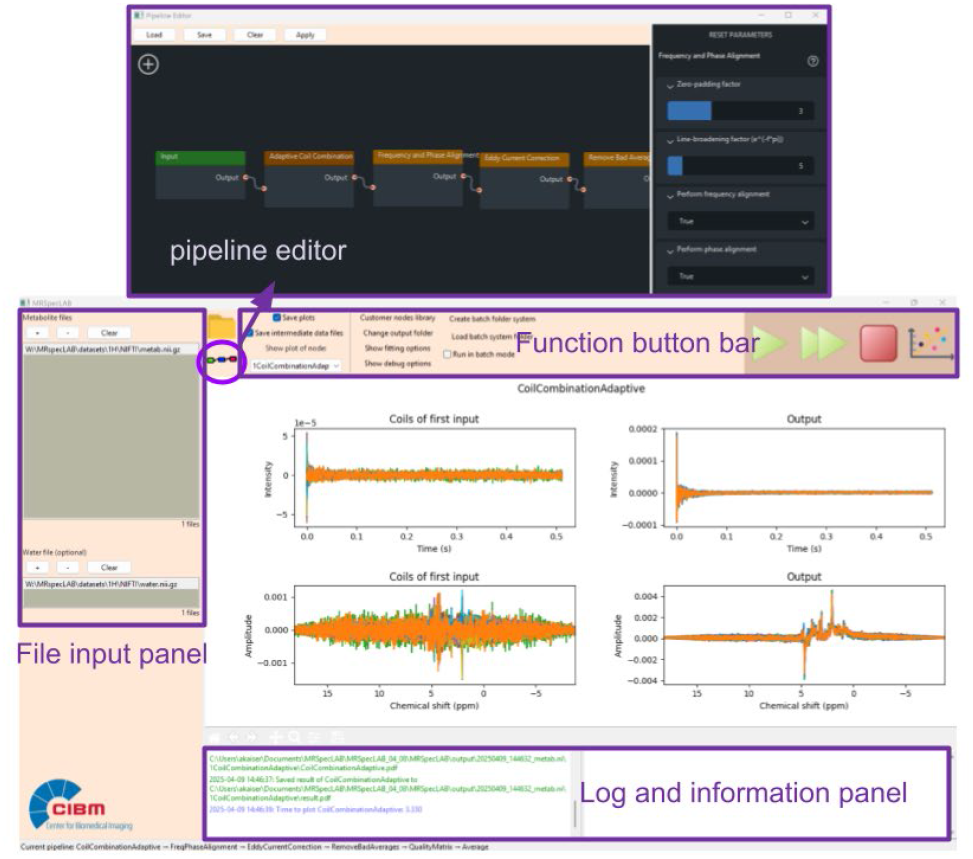
MRspecLAB user interface, consisting of the file import panel (left), function button bar (top), the main plot panel (center), and information panels (bottom), as well as the pipeline editor (the floating window).

The MRspecLAB interface consists of several key components:

- A file import panel on the left side of the window for data input. Data can either be browsed through clicking the ‘+’ button, or by dragging and dropping the files into the panel;
- A function button bar at the top, providing quick access to essential tools like data saving, pipeline execution, and plotting:
  - Open the output directory;
  - Open or change the processing pipeline in the pipeline editor;
  - Two tick boxes for saving intermediate plots and numerical data per processing step;
  - Change the directory of the output files;
  - Fitting options for user defined basis sets, control files, and tissue segmentation files;
  - Show debug options, with a tick box to save error log information;
  - Batch mode setting;
  - Two running modes: single arrow - run the pipeline step-by-step: double arrows - run the pipeline until the end (no in-between plots shown);
  - Stop button to stop any ongoing process;
  - Plotting button to set the display options for MRSI maps.
- A central plot panel where users can view the imported data, intermediate processed plots, fitted spectra, and metabolite maps;
- Two information panels: the bottom-left corner shows real-time updates on processing progress, while the bottom-right corner displays the quantification results extracted from the LCModel coordinate file (.COORD, a text file containing the coordinates of all fitting curves), including quantification results, SNR and FWHM.

The graphic pipeline editor can be accessed from the function button bar, provides users with a fast and intuitive drag-and-drop functionality to build or customize their spectral processing pipelines. By clicking the “+” button within the editor, users can access a library of predefined processing functions (called “nodes” in the following) in the workflow. These nodes can be arranged in any desired order, by dropping them on the canvas and connecting them with already existing nodes. Each node in the pipeline corresponds to a specific processing step. Users can adjust the parameters of each node via the options panel on the right by clicking on the node. The pipeline can be saved as a .pipe file with the ‘‘save’’ button, shared, or stored for future reference, and consequently loaded into the pipeline editor with the ‘‘load’’ button.

### 2.2 Data workflow

MRspecLAB can process MRS and MRSI data in different formats exported from the scanner, or NIfTI format^32^. The software can automatically identify the source and structure of the input data, ensuring compatibility with various data formats, by utilizing suspect^29^. Throughout the workflow, all processed data will be stored and transferred into RAW and NIfTI format. The input data can be processed with either a predefined pipeline, or the pipeline can be adapted to the users’ needs by using nodes in the library, or custom-made nodes. Several predefined pipelines have been included in the current version and introduced in the section below. All example datasets, pipelines, LCModel control files, and basis-set files can be found on Zenodo: https://zenodo.org/records/14866163.

#### 2.2.1 Pre-processing nodes

MRspecLAB is equipped with a set of built-in data processing nodes to handle essential steps in MRS analysis, which are mainly adapted from the existing package^19^ and widely used for MRS and MRSI data processing, mainly including:

1. **Coil combination**: Offers three ways of combining signals from multiple receiver coils, including adaptive combine^33^, S/N^2 34^, SVD^35^.
2. **Frequency and phase alignment**: Corrects frequency drifts and phase variations across sub-spectra.
3. **Apodization**: Applies lorentzian or gaussian apodization functions.
4. **Zero padding**: Extends time-domain signals with zeroes to improve frequency-domain spectral resolution.
5. **Eddy current correction**: Compensates for spectral shape distortion induced by gradient eddy currents.
6. **Bad average removal**: Detects and removes motion-corrupted or outlier sub-spectra.
7. **Quality metrics**: Computes SNR of input metabolite spectra and full width at half maximum (FWHM) of the water peak to enable quick data quality assessment.
8. **Spectral averaging**: Offers three averaging modes, such as overall averaging, blocked averaging, and moving averaging for data quantification.

Detailed information about each processing node and its adjustable parameters is provided in the manual.

#### 2.2.2 Prebuilt pipelines

To support diverse MRS applications, MRspecLAB is equipped with several pre-built pipelines tailored to common applications. These pipelines include:

1. Single-voxel ^1^H MRS (designed for single voxel spectra acquisitions). The provided pipeline includes: adaptive coil combination, frequency and phase correction, eddy current correction, bad average removal, and averaging;
2. Functional MRS (fMRS, supporting dynamic data processing and repeated metabolite quantification for functional studies). The provided pipeline includes: adaptive coil combination, frequency and phase correction, eddy current correction, blocked/moving averaging, and quantification per averaged data set;
3. ^31^P MRS (supports single-voxel ^31^P MR spectra processing). The provided pipeline includes: frequency and phase correction (additional manual frequency and phase correction if necessary), averaging;
4. ^31^P MRSI. The provided pipeline includes: 3D Hanning weighted averaging^36^ and apodization;
5. GABA-edited MRS (MEGA-editing based single-voxel acquisition). The provided pipeline includes: adaptive coil combination, frequency and phase correction, eddy current correction, bad average removal, averaging, and additional manual frequency and phase correction (optional).

#### 2.2.3 Manual frequency adjustment and phasing

The manual frequency adjustment and phasing tool provides an easy way to fine-tune processed data before fitting. After completing the pipeline before quantification, users can access the manual frequency and phase adjustment option via a popup window. In this panel, they can adjust the spectral frequency offset by entering a value or using the slider to shift the entire spectrum along the frequency axis. This is an optional fine-tuning step that allows users to align peaks with their expected positions, ensuring the spectrum is correctly centered for further analysis.

Users can also manually input values for zero-order and first-order phasing. As users make adjustments, the updated spectrum is displayed in real-time in the plot panel, providing immediate visual feedback. Once the adjustments are complete, these settings will be applied to the averaged data before moving on to the fitting stage.

#### 2.2.4 Spectral fitting

Spectral fitting in MRspecLAB is realized with LCModel Version 6.3. For LCModel-based spectral fitting, a basis-set file (a collection of simulated or experimentally acquired model metabolite spectra compatible with the input data) and a control file (a text file specifying the spectral fitting parameters, and options for spectral decomposition and individual metabolite concentration estimation) are needed. MRspecLAB will attempt to identify a default basis set by matching the sequence parameters extracted from the input data header, as well as the LCModel control file (this list will be lively updated and expanded in the folders of https://github.com/MRSEPFL/MRSprocessing/basissets & https://github.com/MRSEPFL/MRSprocessing/controlfiles). Alternatively, users can supply their own basis set and LCModel control file via the “fitting options” button on the top panel before running the pipeline analysis. Once provided, the software automatically directs the processed data to the fitting procedure at the end of the pipeline.

Additionally, users can provide grey matter (GM), white matter (WM), and cerebral spinal fluid (CSF) probabilistic tissue segmentation files (probabilities between 0-1) to correct metabolite concentrations for water content differences, which improves quantification accuracy by accounting for voxel composition and tissue-specific relaxation effects. For this, any segmentation program can be used, which can segment a T1-weighted anatomical scan into these three tissue components. The user can upload the tissue segmentations in the appropriate fields through the “fitting options”. The voxel-specific fractions will then be calculated and used to determine the water concentrations within the voxel^37^. The water content value will be automatically adapted into the control file for metabolite concentration analysis with LCModel.

#### 2.2.5 Output structure

Clicking the folder icon in the function bar opens the designated output folder, where users can access intermediate results, processed data, and LCModel fitting outcomes. The output of the autorun analysis is systematically organized within a default output directory, though users can select a custom directory using the “change output folder” button. If the “save intermediate data files” option is selected, the processed raw data will be saved as ASCII files (.RAW) and NIfTI-MRS files (.nii). The processing pipeline consists of multiple sequential steps; The results of each step are stored in a dedicated subfolder, containing processed spectra, and diagnostic plots (.PDF). LCModel fitting results are saved in a separate LCModel folder. Additionally, each run generates an MRSinMRS table^3^ based on REMY tool^4^, a standardized format for reporting MRS acquisition parameters, and processing procedures, ensuring consistency and reproducibility in research. Finally, a .pipe file will be created, which saves the pipeline used in the current processing. Details about the output files and its structure are provided in the Supplement 8.

#### 2.2.6 Self-defined processing nodes

To create a custom node that can be included into the pipeline, a developer can write a Python class that defines the functionality of the node. This class should follow the predefined framework provided by MRspecLAB, specifying the adjustable parameters, and the processing. This file should be placed in the designated folder within the MRspecLAB directory, which can be visible in the “Custom node library” on the function bars after re-launching MRspecLAB. To use a custom node, open the pipeline editor, and the new node can be dragged and dropped into the canvas as prebuilt nodes, and connected with the other nodes. Users can then adjust the node’s parameters (if configured) as needed.

### 2.3 Batch processing

MRspecLAB now features a batch processing mode to facilitate efficient analysis of multiple MRS datasets. This mode includes three key functionalities:

1. **Make batch folder system**
  ∘ This function creates a structured batch folder system in a user-specified directory.
  ∘ The user is prompted to enter the **study name** and define the **number of participants**. The generated folder structure follows this format:

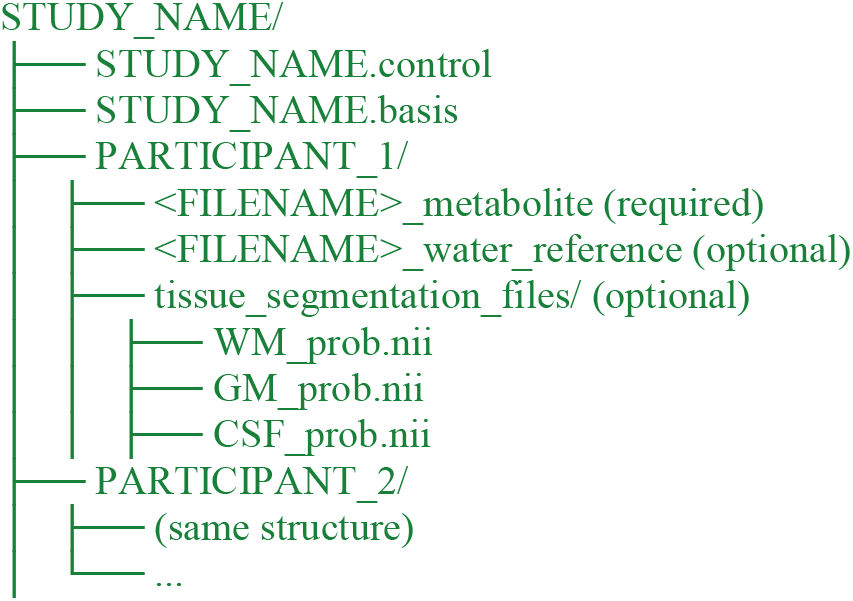
  ∘ The .control and .basis files, required for processing, are placed in the main study folder.
  ∘ The user is responsible for manually placing the necessary files into the corresponding participant folders.
2. **Load batch folder system**
  ∘ This function allows users to load an existing batch folder structure for processing.
  ∘ The tool automatically checks whether the required files (.control, .basis, and metabolite files) are present.
  ∘ It also verifies that the provided metabolite file types match the supported input formats (as specified in the methods section).
  ∘ Missing or incorrectly placed files trigger warnings.
3. **Run batch mode (checkbox option)**
  ∘ Enabling this option allows MRspecLAB to automatically process all participants within the loaded batch folder structure.
  ∘ The tool sequentially applies the same processing pipeline to each participant, ensuring consistency.
  ∘ If optional files (e.g., tissue segmentation files or water references) are provided, they are included in the workflow.
  ∘ Any errors, missing files, or incompatibilities are flagged before processing begins to ensure data integrity.

## 3 Results

We demonstrate the provided pre-defined pipelines along with example datasets in detail. The .pipe files, .control files, and basis sets along with example datasets are available on the Zenodo repository: https://zenodo.org/records/14866163.

### 3.1 Application 1: Processing and quantification of single voxel ^1^H MRS data

Here, we demonstrate the workflow for single-voxel spectroscopy (svs) ^1^H MRS data processing.

#### 3.1.1 Data input

The example dataset included water-suppressed data acquired using the STEAM sequence (TE/TR = 4.5/4000 ms, bandwidth = 4 kHz, voxel size = 30 × 30 × 30 mm^3^, 32 averages) and unsuppressed water data obtained using the same sequence and parameters, from the human parietal lobe on a Siemens Terra.X 7T scanner with a 1TX / 32RX Head Coil (Nova Medical). The input data were both in Siemens raw data format (.dat).

#### 3.1.2 Spectral processing pipeline construction and outcome

Here the default pipeline as described above was used. The first step involved adaptive coil combination, as the raw data consisted of individual signals from multiple channels, optimizing the SNR based on the water reference data. After coil combination, the data underwent the alignment of individual transients within the dataset. The frequency and phase alignment procedure uses the N-acetylaspartate (NAA) peak at 2.02 ppm as the reference for short-TE ^1^H MRS data. Next, eddy current correction was performed using the water reference spectra to correct for eddy current artifacts. Next, the similarity of each transient was assessed using the root-mean-squared (RMS) value of its difference from the mean of all transients, and any outliers were deleted. Finally, all remaining transients were averaged together and sent to LCModel for fitting and spectral quantification.

Figure 3 illustrates the workflow for executing the defined pipeline to obtain the quantification results. A more detailed demonstration of this application case, including step-by-step processing and visual outputs, can be found in Supplement 1. An example control file for short-TE svs ^1^H MR spectral fitting is provided in Supplement 2.

**Figure 2:**
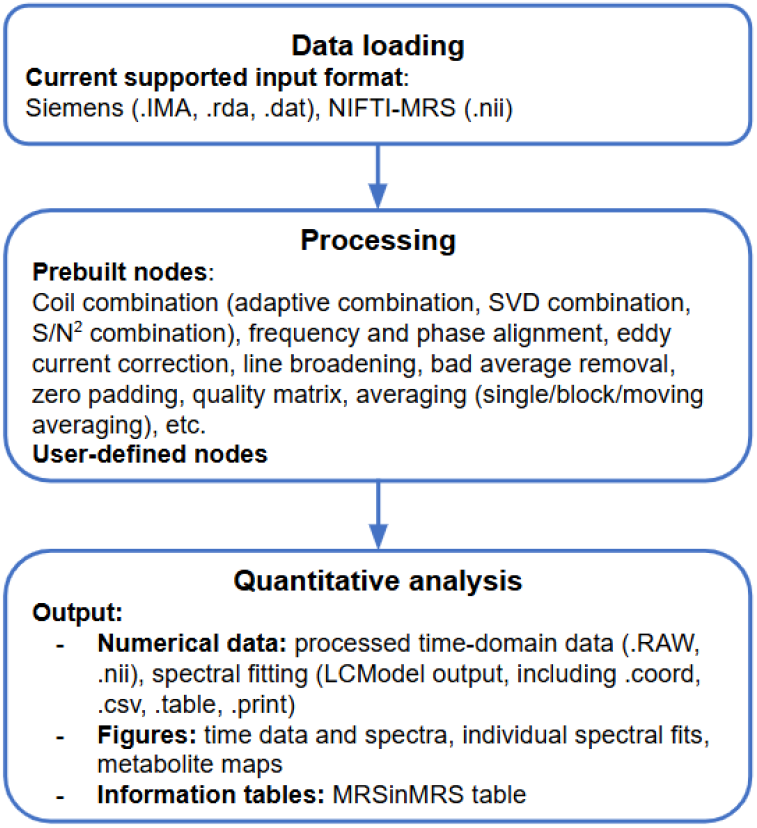
MRspecLAB data workflow: from raw data to processed data and quantification results.

**Figure 3:**
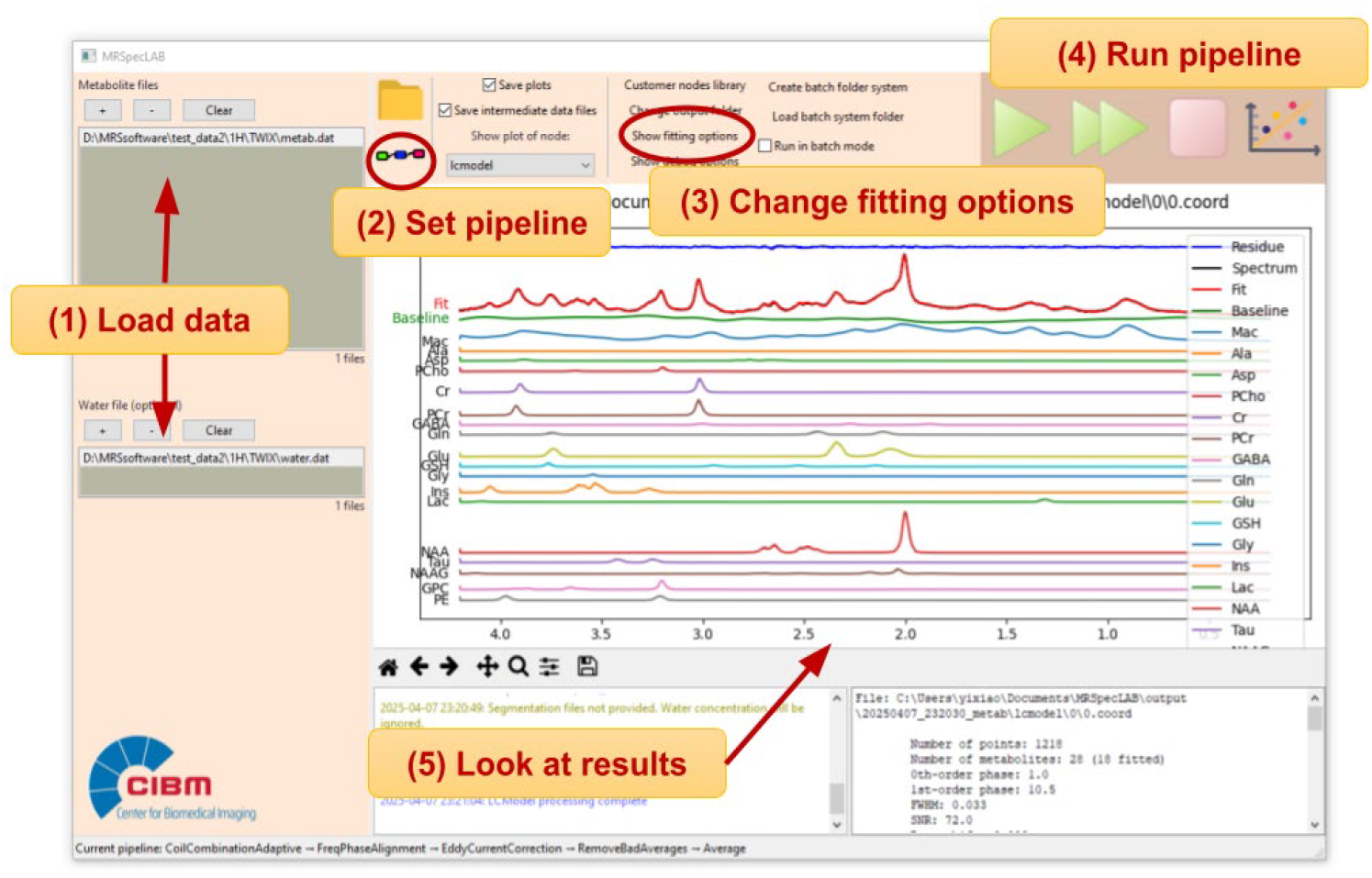
Workflow of using MRspecLAB to process 1H MRS data and obtain quantification results. Users follow these steps: (1) Load the water-suppressed data in the top section and the unsuppressed water data in the bottom section. (2) Open the pipeline editor to load, review or modify the processing pipeline as needed. (3) Change the fitting options for LCModel if desired: Add a .basis set, .control file (Supplement 2), tissue segmentation files. (4) Click the “run step-by-step” or “run” button to execute the pipeline and perform spectral fitting. (5) View the fitted individual spectra in the central plot panel and examine the numerical results in the information panel.

### 3.2 Application 2: Processing and quantification of fMRS data

The fMRS processing pipeline builds on the processing steps detailed in Application 1, and introduces the spectral averaging techniques. Spectral averaging is critical in fMRS data processing, as it enhances the SNR, which is essential for detecting the subtle metabolic changes associated with functional brain activity. MRspecLAB provides several flexible averaging options, such as block averaging, and moving averaging, enabling users to tailor the process to their specific experimental conditions.

#### 3.2.1 Data input

^1^H-MR spectra were acquired using semi-adiabatic spin-echo full-intensity-acquired localization (sSPECIAL)^38^ (TR/TE = 4000/16 ms, bandwidth = 4 kHz, 4096 data points) from the dmACC (128 measurements, 16 per block, voxel size = 20 × 20 × 25 mm^3^) on a 7T Siemens Terra.X MR system (Siemens Medical Solutions, Erlangen, Germany) using 1TX / 32RX Head Coil (Nova Medical). The commonly used symbol digit matching task (SDMT) was utilized in the MRI^39^. During repeated MRS scans, participants were presented with a table of numbers and their corresponding symbols. The participants were then asked to match a novel set of symbols to their associated numbers in the active condition. In the rest condition, the table was empty and participants were instructed to read the number on the screen. These conditions were presented in 8 blocks (4 active, 4 rest), each 128 seconds, with 64 stimuli randomly presented for 2 seconds each.

To validate the functionality and highlight the effectiveness of the averaging techniques, we processed this example dataset using both blocked and moving averaging methods. The detailed information and output files can be found in Supplement 3. By serving as a practical reference, these results guide users in applying and optimizing these techniques for their own datasets.

#### 3.2.2 Blocked averaging

In contrast to overall averaging used in Application 1, where all transients are averaged together to maximize SNR for a single quantification result across the entire dataset, MRspecLAB also supports block-based averaging, which divides the data into smaller time segments and averages within each block, allowing users to study metabolic changes over specific time windows, such as rest and active states.

For block averaging, MRspecLAB provides three key parameters to control the process, as shown in the right panel of Figure 4: (1) number of measurements in an experimental block: This defines the original number of transients within each block of raw data, representing the unaveraged input for that block; (2) number of averages to produce per block: This specifies the desired number of averaged spectra per block. For example, if a block contains 16 transients, setting this parameter to 4 will produce 4 averaged spectra by averaging every 4 transients together; (3) number of block types: this determines the distinct experimental conditions (e.g., rest vs. active) to be processed separately. Each block type is treated independently.

**Figure 4:**
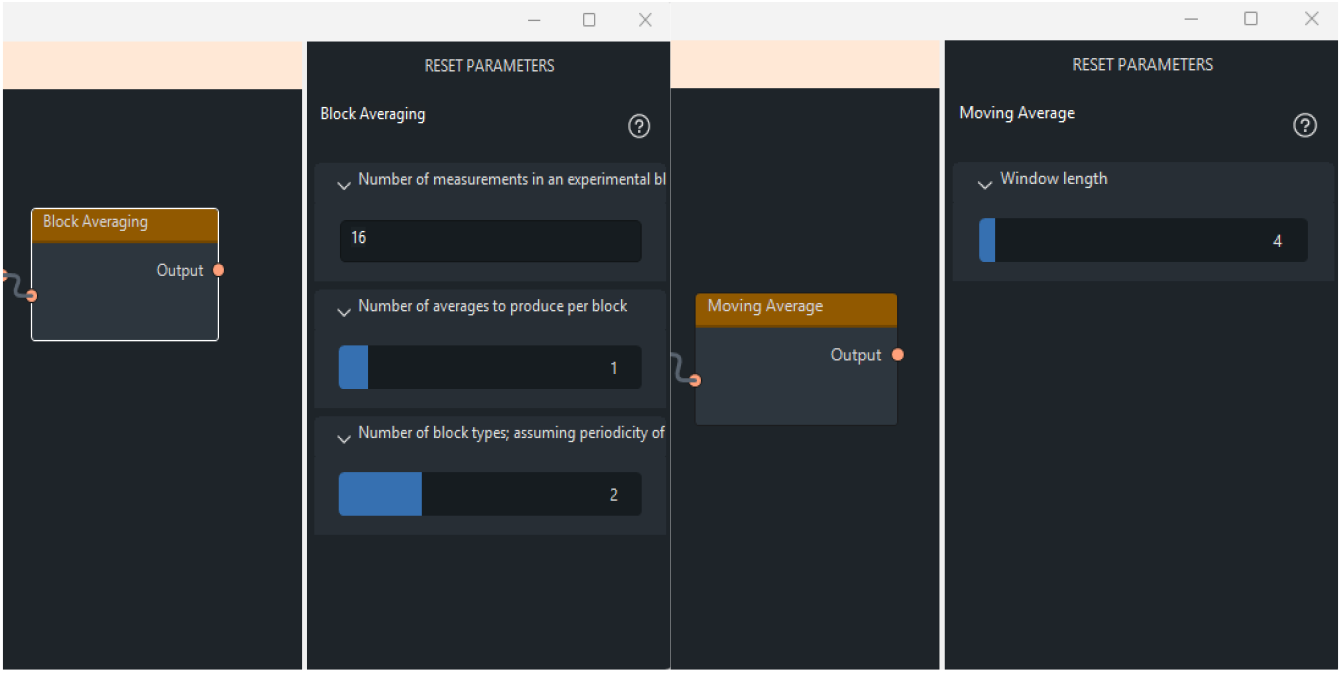
Integrating block/moving averaging into the processing pipeline for fMRS. On the right panel, the user can adjust several parameters of the function, such as the number of measurements in an experimental block, the number of averages per block, and the number of block types. A) In this dataset, two block types (rest and active) were considered, with each state including 16 transients. To generate the averaged dataset, 1 average per block was generated, resulting in 8 blocks for fitting. B) The window length of 4 is defined to average every 4 consecutive transients together, with the window advancing by 1 transient at each step.

The block averaging process involves grouping the transients within each block based on the defined number of measurements. Transients within each block are averaged according to the specified number of averages per block, effectively reducing the total number of spectra while enhancing SNR. Users can tune this parameter and try to make a balance between data quality and temporal resolution for experiments with multiple experimental conditions.

After the averaging process, each averaged spectrum was fitted and quantified. The software automatically generates outputs for each averaged spectrum, including metabolite concentrations, fit diagnostics, and spectral fits. These results are organized into distinct directories corresponding to the block types and averaging configurations, enabling users to easily access and analyze condition-specific results.

#### 3.2.3 Moving averaging

For experiments requiring dynamic quantification, such as task-related fMRS studies or if dynamics within blocks want to be investigated, a “moving averaging” node is available. This approach calculates the average over a sliding window, maintaining temporal resolution while improving SNR. This is particularly useful for capturing transient metabolic fluctuations without sacrificing data quality.

For moving averaging, MRspecLAB provides a parameter called “window length,” which defines the number of transients within the sliding window used for averaging, as shown in Figure 4. For instance, setting a window length of 5 means that every 5 consecutive transients are averaged together, with the window advancing by 1 transient at each step. This method applies the moving average by sliding the defined kernel across the dataset, averaging the transients within the window at each step. The process is repeated until the entire dataset is processed, producing a series of averaged spectra. As with blocked averaging, LCModel is employed to analyze each spectrum generated through the moving averaging process.

#### 3.2.4 Comparison of spectral quality metrics between MRspecLAB and FID-A

To assess the consistency between MRspecLAB and other existing toolboxes in processing MRS data, spectral quality metrics—SNR and water linewidth, were evaluated for five participants’ data. The dataset includes a short pre-acquisition of 8 MRS measurements acquired under the protocol described in Application 2. The raw input data were identical for both tools, provided in Siemens TWIX format, and underwent the same preprocessing steps: water-signal-based coil combination, frequency and phase alignment, eddy current correction, outlier average removal, and signal averaging.

Figure 5 illustrates the correlation between the two tools for SNR and water linewidth with the described datasets. The SNR was computed based on the peak height of the NAA signal at 2.02 ppm divided by the RMS noise in the 0.2 to 0.5 ppm region, while water linewidth (FWHM of the water peak) was measured in Hz. A strong linear relationship was observed for both metrics, with data points closely aligned along the line of identity (x = y), indicating high agreement. Statistical analysis revealed significant correlations (r(3)=0.92, p=0.03; r(3)=0.95, p=0.01), confirming a significant and consistent relationship between the outputs of the two toolboxes.

**Figure 5:**
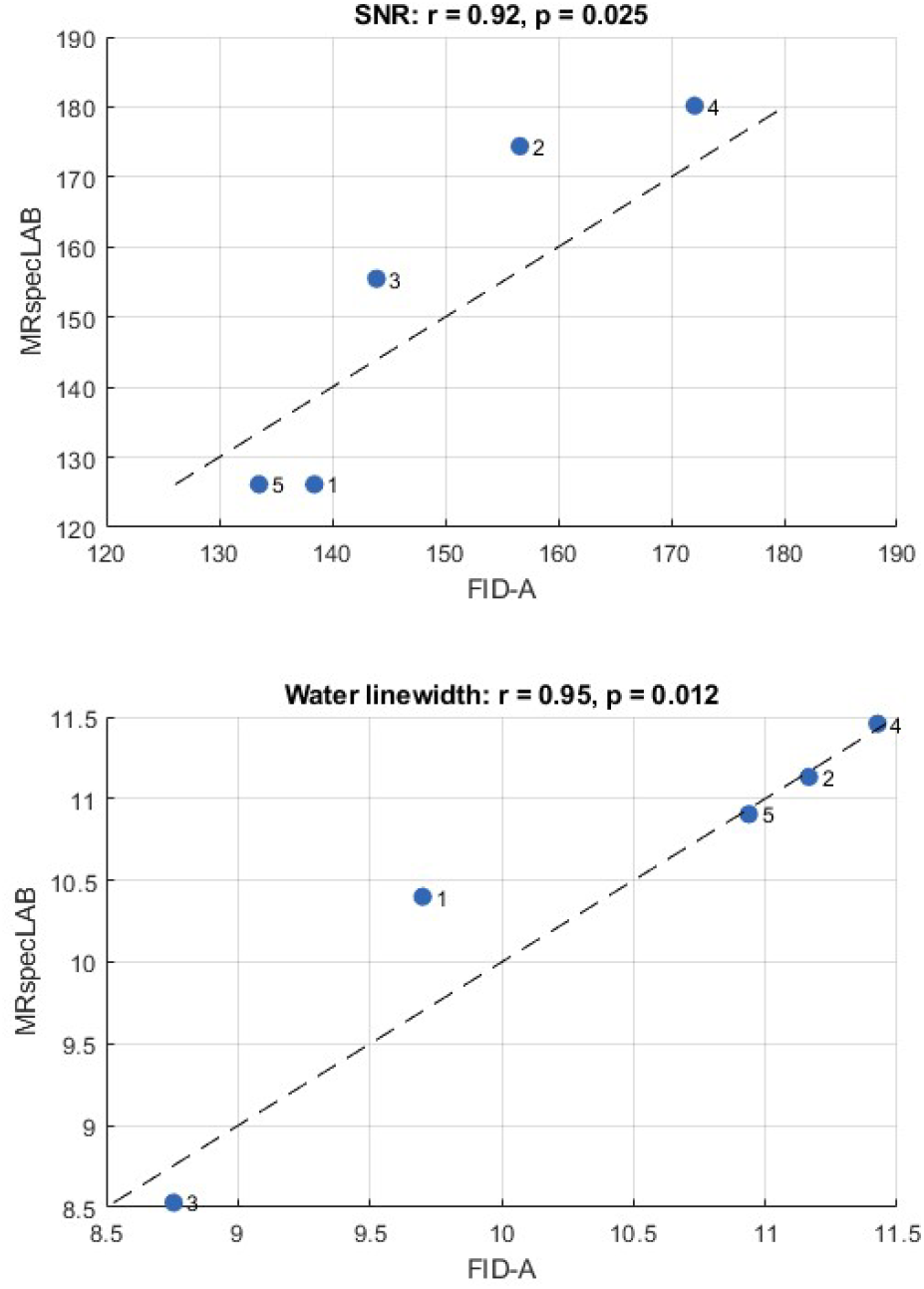
Correlation between post-processed spectral quality metrics (SNR and water linewidth) obtained from FID-A and MRspecLAB for five participants. The input data consisted of a short pre-acquisition of 8 measurements for each participant, acquired using the protocol outlined in Application 2. Identical raw data in TWIX format were processed by both FID-A and MRspecLAB, following the same preprocessing pipeline: coil combination based on the water signal, frequency and phase alignment, bad average removal, eddy current correction, and signal averaging. SNR was calculated from the NAA peak at 2.02 ppm divided by the RMS noise from 0.2–0.5 ppm, and the linewidth (in Hz) was measured from processed water spectra for all 5 participants. Each data point represents one participant and is labeled accordingly. The dashed line indicates the line of identity (x = y). Pearson correlation coefficients (r) and corresponding p-values (p) are provided, demonstrating strong agreement between FID-A and MRspecLAB in post-processing spectral quality.

Additionally, detailed comparisons of the individual post-processed spectra, and quantification results for major metabolites (e.g., tNAA, tCr, Glu), are presented in Supplement 3.

### 3.3 Application 3: single-voxel ^31^P MRS data processing

In addition to supporting ^1^H MRS data, MRspecLAB is also equipped to process X-nuclei data, including ^31^P MR spectra.

#### 3.3.1 Data Input

MR experiments were performed on a 7T/68 cm MR scanner (Siemens Medical Solutions, Erlangen, Germany) with an in-house-built ^1^H quadrature surface coil (10-cm diameter) and a single-loop ^31^P coil (7-cm diameter) for the coverage of the human occipital lobe. B0 field inhomogeneity was optimized in a voxel of interest (VOI) (50×30×40 mm^3^). Localized ^31^P MR spectra were acquired using a 3D-ISIS sequence (TR/TE = 3000/0.35 ms, voxel-size = 55 × 20 × 25 mm^3^, averages/block=16/6, bandwidth = 6 kHz, number of points = 2048).

The processing pipeline used was described above. A surface coil was utilized in this experiment, so no coil combination was needed. 31P frequency and phase correction were applied. The individual spectral fitting can be obtained in the end as shown in Figure 6. Supplement 5 provides a more detailed demonstration of this application case, including step-by-step processing and visual outputs.

**Figure 6:**
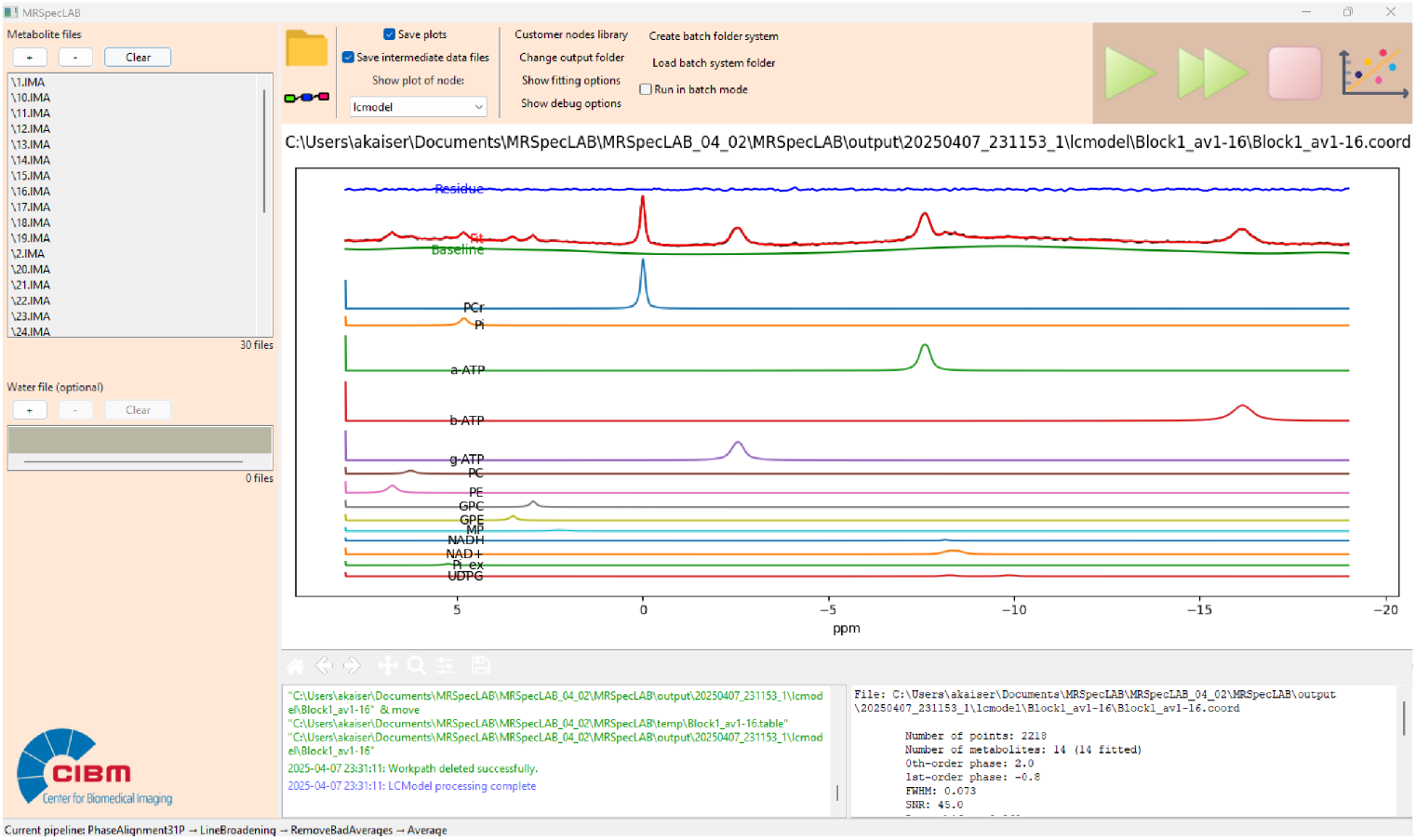
Example fitting result for a single-voxel 31P MR spectra (3D-ISIS, TR/TE = 3000/0.35 ms, voxel size = 55 × 20 × 25 mm3, averages/block=16/6, bandwidth = 6 kHz, number of points = 2048).

### 3.4 Application 4: ^31^P MRSI data processing and metabolite mapping

MRspecLAB also provides the plotting tools specifically designed for visualizing the concentration spatial distribution in multivoxel data. Here is an example for processing, quantifying, and visualizing the ^31^P CSI-FID data.

#### 3.4.1 Data input, processing, and fitting

The input data is acquired by a ^31^P CSI-FID sequence with the following parameters: FOV = 200 × 200 × 80 mm^3^, matrix size = 16 × 16 × 8, TE/TR = 2.3/260 ms, flip angle = 33°, bandwidth = 6 kHz, vector size = 1024, 160 averages. The dataset is in Siemens .rda format, and is processed by applying the Hanning filter using the node named 3D Hanning filter (window size = 16 × 16 × 8), the phase correction, and the line broadening CSI (Gaussian, 5 Hz) to the averaged spectra to enhance the SNR. The step-by-step processing, visual outputs, and fitting, can be found in Supplement 6.

The ^31^P MR spectra are subsequently analyzed by LCModel on a voxel-by-voxel basis using the provided basis set. This basis set includes key phosphorus-containing compounds such as phosphocreatine (PCr), intracellular and extracellular inorganic phosphate (Pi), diphosphates (NAD, UDPG), triphosphates (α-, β-, and γ-ATP), phosphoethanolamine (PE), phosphocholine (PC), glycerophosphoethanolamine (GPE), and glycerophosphocholine (GPC). The basis set and the control files used in the fitting process can be found on the Zenodo repository. For each voxel, the output is stored in a dedicated subfolder named according to its 3D spatial coordinates.

#### 3.4.2 Metabolite concentration map visualization

MRspecLAB has a built-in plotting tool designed for generating metabolite maps, which can be accessed via the plotting button located in the top right corner of the panel. Clicking on the plotting button can launch the visualization parameter interface (Figure 6). By using this tool, users can visualize the spatial distribution of metabolite concentrations once the output data is available.

A selection list will display the available metabolites, allowing users to choose one for generating a corresponding 2D spatial concentration map. Users can specify the map orientation and select the slice number they wish to visualize. Additionally, there is an option to choose a reference metabolite. When a reference is selected, only pixels where the CRLB (Cramér-Rao Lower Bound) values for both the reference and selected metabolites fall below the threshold will be displayed, ensuring reliable data visualization. Furthermore, once a reference metabolite is chosen, users can opt to display a relative concentration map, which represents the ratio of the selected metabolite to the reference metabolite. To enhance visualization, users can also customize the colorbar scale, adjusting it for optimal contrast and clarity in the displayed maps.

Moreover, the brain masks can be calculated from the corresponding anatomical images. Here, the corresponding 3D ^1^H anatomical images acquired using a GRE sequence (TE/TR = 2.82/6.50 ms, α = 4°, 1 mm^3^ isotropic resolution) were loaded. After selecting the orientation of the anatomical image and the corresponding slice number, the plotting tool can generate a brain mask and apply it to the map based on the chosen image. Figure 7 illustrates one slice of the NAD^+^ maps derived from this workflow.

**Figure 7:**
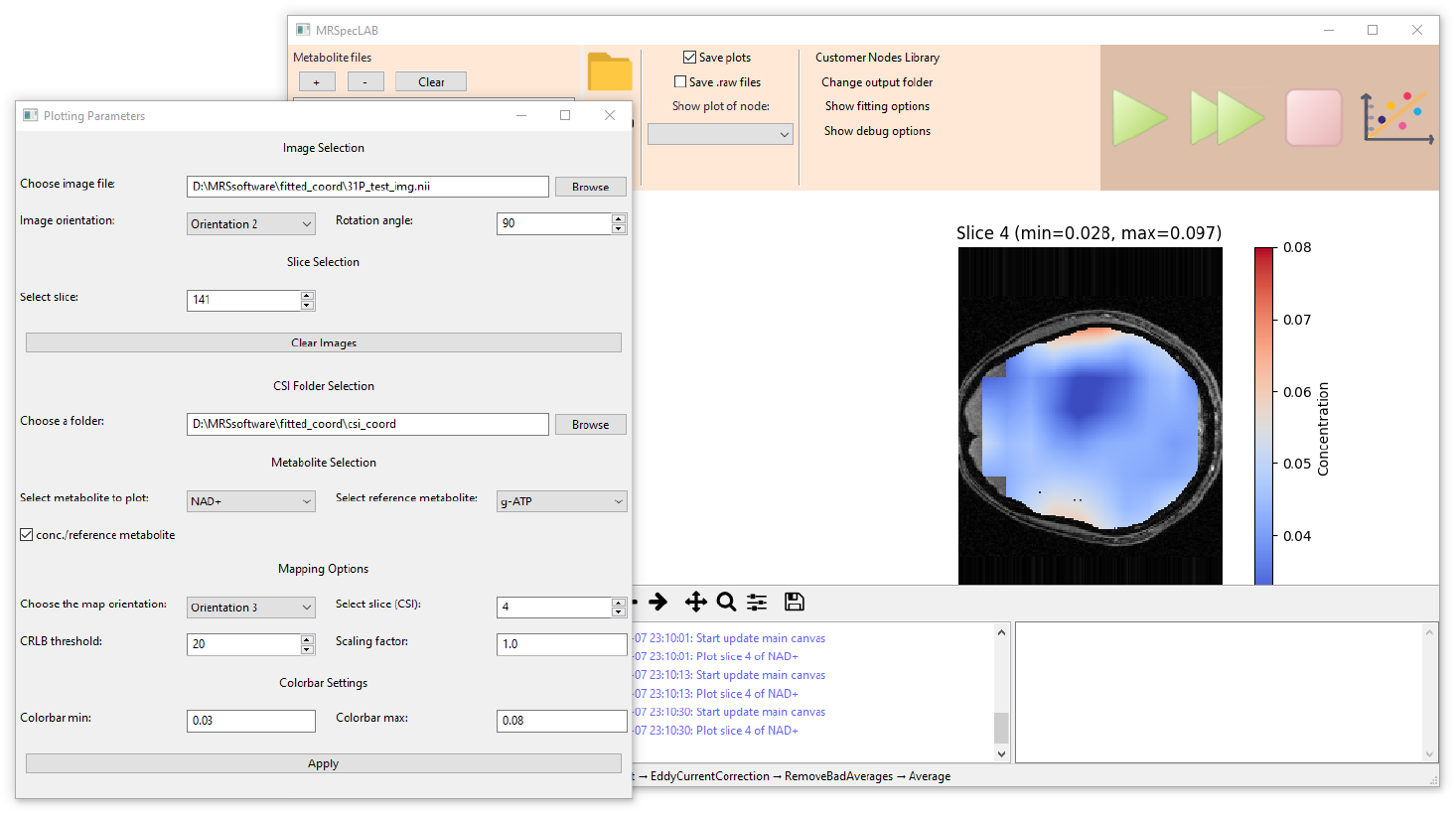
Application for processing and quantifying ^31^P CSI-FID data (rda format, FOV = 200 × 200 × 80 mm3, matrix size = 16 × 16 × 8, TE/TR = 2.3/260 ms, flip angle = 33°, bandwidth = 6 kHz, vector size = 1024, 160 averages). Corresponding 3D ^1^H anatomical images were acquired using a GRE sequence (TE/TR = 2.82/6.50 ms, α = 4°, 1 mm3 isotropic resolution). The selected 2D concentration maps can be visualized using the implemented plotting tool based on spectral fitting results.

### 3.5 Application 5: Spectral-editing for GABA measurement

Mescher–Garwood (MEGA) J-difference editing^40-42^ is a common method used for the detection of the resolved GABA signal at 3.0 ppm, which is usually difficult to measure due to its overlap with other intensive metabolite resonances. The processing of MEGA data often requires subtraction of two sub-spectra to obtain the resolved GABA signal. In MRspecLAB, the MEGA data is handled the same way as svs ^1^H data, given that the combination of sub-spectra in the sequence is direct averaging. The alignment between edit-on and edit-off spectra is critical in MEGA-editing to obtain a resolved GABA peak.

#### 3.5.1 Data input

Here, we processed an example dataset acquired using MEGA-sSPECIAL sequence^43^ with the following parameters: TE/TR = 80/4000 ms, bandwidth = 4 kHz, voxel size = 30 × 30 × 20 mm^3^, 32 acquisitions with 4 sub-spectra each (two additional scans are for 1D-ISIS module in the sSPECIAL).

#### 3.5.2 Processing and quantification

The processing steps follow a similar set to those in Application 1, including adaptive coil combination, frequency and phase correction, eddy current correction, bad average removal, and averaging. One example of the fitting result is shown in Figure 8.

**Figure 8:**
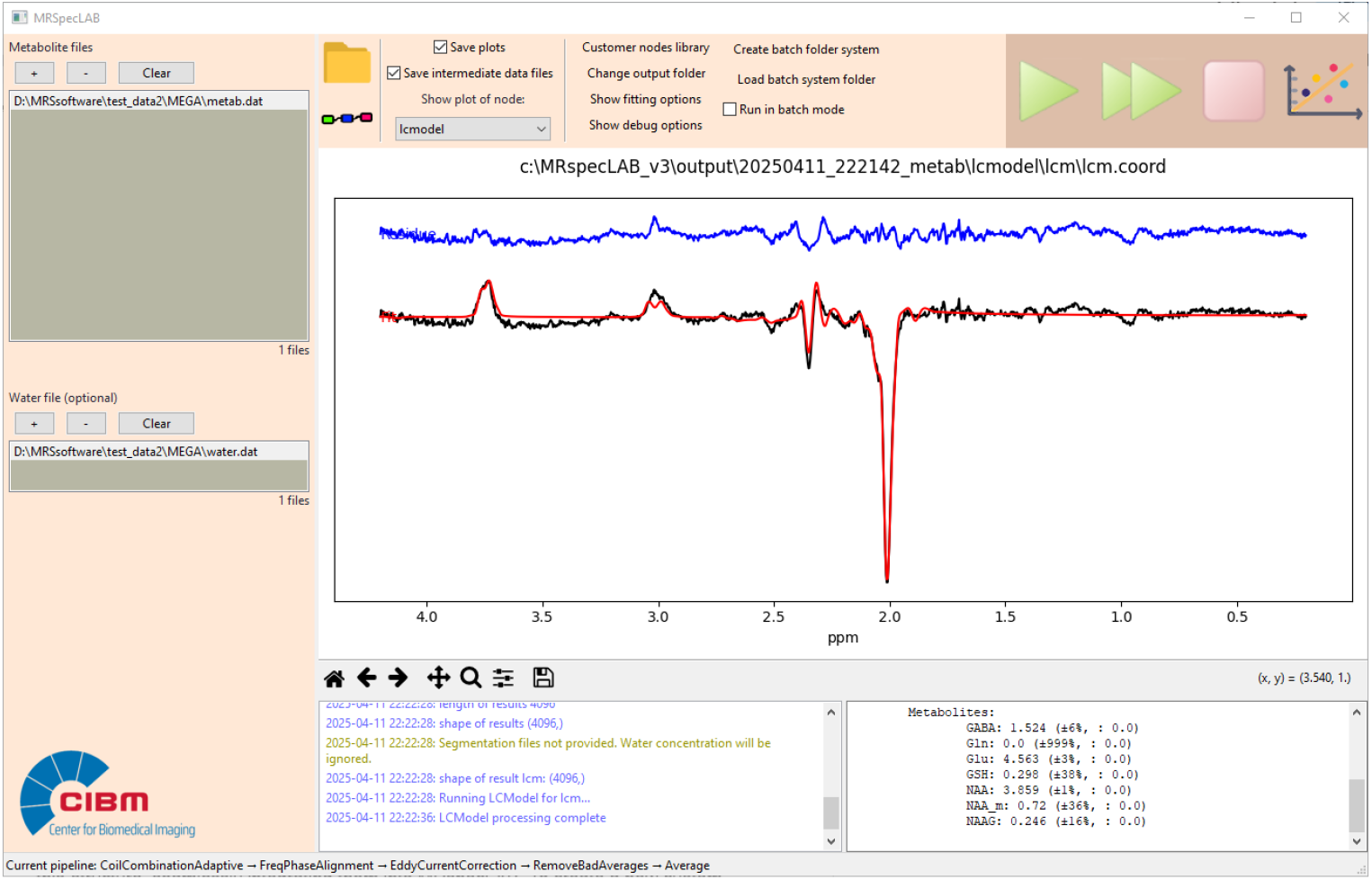
Example fitting result for an edited GABA sequence (MEGA-sSPECIAL, TE/TR = 80/4000 ms, bandwidth = 4 kHz, voxel size = 30 × 30 × 20 mm^3^, 32 acquisitions).

### 3.6 Application 6: Development of custom processing nodes and self-defined pipelines

Each custom node should be implemented as a Python class, adhering to a predefined framework. This framework includes essential components such as the basic information (e.g., label, author, and description), parameter definitions, and core processing functionality. Developers can develop and share new nodes using this structure, seamlessly integrating them into MRspecLAB. To create a new custom node, follow these steps (Figure 9): (1) node’s information definition: provide metadata such as the node’s label, author, and description to identify and describe the node within the pipeline editor; (2) key parameter definition: specify adjustable parameters that will appear in the right panel when the node is selected in the pipeline editor; (3) processing function: implement the process method to define how the node processes input data; (4) visualization (optional): Implement the plot method to generate diagnostic plots of both input and processed data. These plots will be displayed in the main window of the software upon completion of the processing step; (5) register the node: use api.RegisterNode to add the node to the library, making it accessible via the GUI.

**Figure 9:**
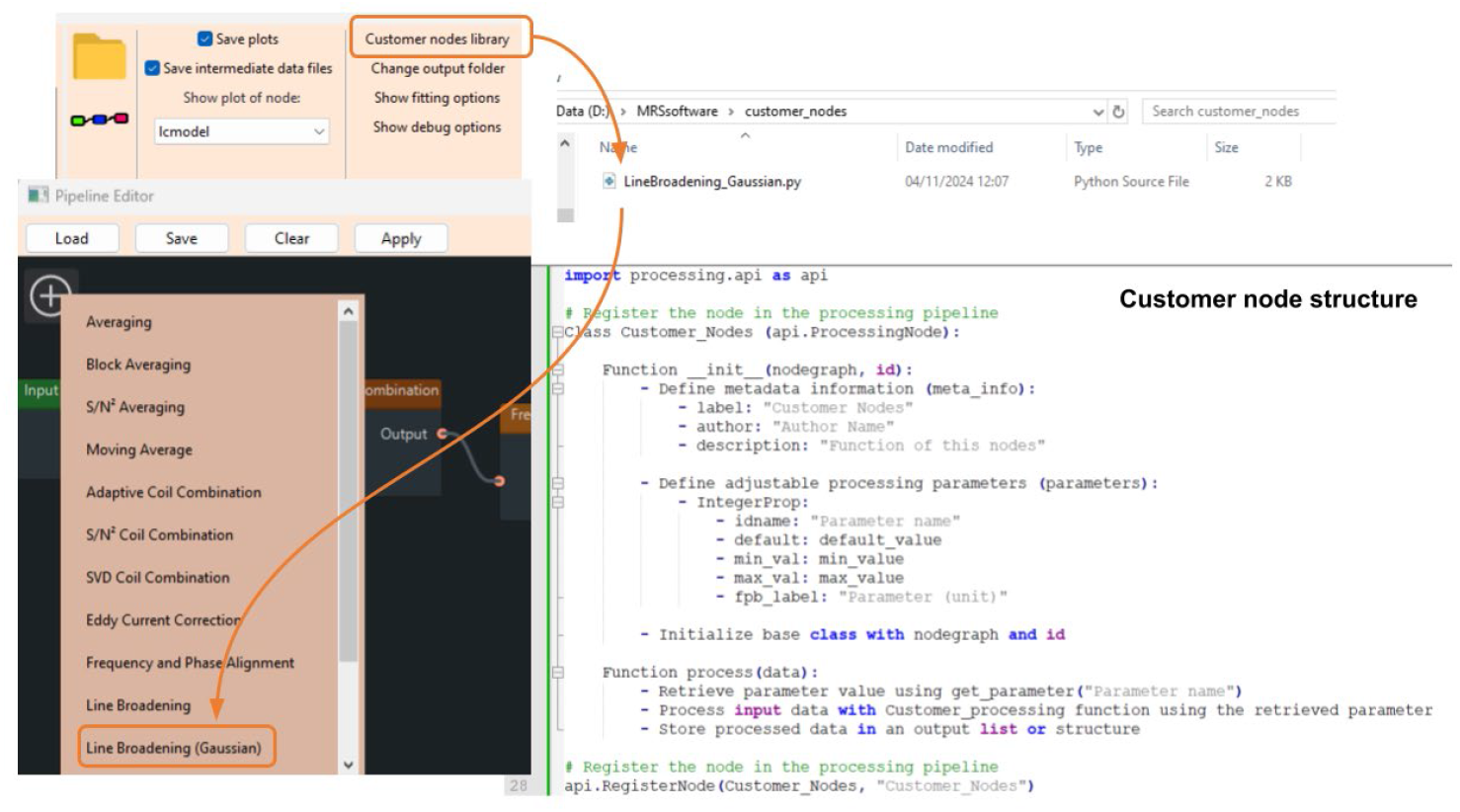
Adding custom nodes. In the main window, users can select a custom node library, which loads all available custom nodes within the chosen directory. The new nodes appear on the list with the prebuilt nodes and can be easily added to the pipeline diagram using a drag-and-drop action. Each custom node is structured as a Python class, with a predefined framework provided.

An example code for creating a custom node can be found in Supplement 7. For users seeking to utilize a pipeline beyond the prebuilt options, the first step is to load all required external processing node scripts into the custom node library. After relaunching, open the pipeline editor and load an existing .pipe file to configure a specific pipeline. Additionally, a GitHub repository is available where all existing nodes are stored, and developers can contribute their nodes via push requests. Submitted nodes undergo validation before being added to the library for non-developer users.

## 4 Discussion

MRspectLAB serves as a collaborative platform that bridges the gap between the MRS technical experts, neuroscientists, and clinicians, supporting the interdisciplinary collaboration between those focused on the technical aspects of MRS and those applying it in clinical or neuroscientific contexts.

Its modular design empowers users to create specific processing pipelines, where each processing step is defined as an independent node that can be easily tuned and rearranged (drag-drop-connect) in a graphic interface to meet the needs of the specific datasets. The user-developed nodes can be shared across the research community and seamlessly integrated into the graphical pipeline editor by others. This allows users to focus their efforts on refining and optimizing data processing workflows, rather than spending time navigating or modifying complex code.

MRspecLAB can be freely downloaded as a compiled executable, and can be run directly on Windows without the need for any installation steps or environment configuration. It supports a wide range of data formats from vendors and software versions. It also supports the NIfTI-MRS format^28^, which has been proposed as a standard spectroscopy data format, eliminating the need for manual format conversions. Data processed in MRspecLAB can be exported in ASCII and NIfTI formats compatible with other widely used packages, ensuring compatibility with other software packages, and thereby allowing researchers to integrate their analyses with other established tools.

MRspecLAB offers an intuitive interface and graphical drag-and-drop pipeline editor, making MRS/MRSI research accessible to a broad user base. Even users with minimal experience in MRS/MRSI data processing can easily analyze their data. Its user-friendly design provides a streamlined workflow, enabling students, clinicians, and researchers to engage with MRS/MRSI data analysis efficiently. With the potential of supporting a comprehensive range of spectral processing tasks, including X-nuclei datasets, MRspecLAB is well-suited for both routine and advanced MRS/MRSI research. The pipeline files together with the data can be shared with publications to facilitate research reproducibility.

Processing and quantification of MRS data by MRspecLAB has been validated by comparison with FID-A combined with LCModel. The results indicate that MRspecLAB achieves comparable metabolite concentration estimates and spectral quality metrics, supporting the reliability for automated spectral preprocessing using MRspecLAB. Predefined pipelines in MRspecLAB have been optimized based on expert-recommended workflows^18^, ensuring accurate and reproducible analysis.

A key strength of MRspecLAB is its modular and adaptable design. Its architecture allows users to customize workflows to suit their specific datasets, with each processing step defined as an independent node that can be easily rearranged and adjusted. Users can also develop and integrate new nodes within the provided framework, facilitating expansion without requiring extensive programming skills. Additionally, custom modules and pipelines can be shared within the research community, fostering innovation and encouraging collaborative development of advanced MRS/MRSI processing methods.

One limitation might be the reliance on LCModel for spectral fitting, while robust, may limit flexibility for users who wish to employ alternative quantification methods. Incorporating plug-ins to interface with other popular software like FSL, SPM, Osprey may be a direction for future development. However, MRspecLAB allows to save the intermediate processed data in .nii format, which could be read into any other fitting program afterwards, if needed, and additionally, fitting algorithms could be implemented as a customized node. As an open-source platform, MRspecLAB continues to evolve based on user feedback and contributions from the research community. Future development plans include integrating additional processing nodes and expanding support for more experimental modalities, such as ^13^C, and ^2^H MRS and ^31^P MR fingerprinting. Community-driven enhancements will play a vital role in extending the software’s capabilities, with MRS researchers contributing new nodes, sharing processing pipelines, and neuroscientists and clinical scientists offering usability feedback.

## 5 Conclusion

MRspecLAB is a collaborative platform for MRS processing, designed to bridge the gap between technical experts and application-driven users in MRS/MRSI research, clinical applications, and education. Its intuitive graphical interface and modular pipeline editor enable experienced users to develop and share advanced functions while allowing other users to seamlessly integrate them into their workflows. With robust support for both proton and X-nuclei single-voxel and MRSI datasets, MRspecLAB serves as a dynamic platform for knowledge exchange, innovation, the advancement of MRS methodologies, bridging MRS methods with clinical applications and enhancing the value of MRS in clinical and neuroscience research.

## 6 Data Availability Statement

The code and manual are openly available on GitHub (https://github.com/MRSEPFL/MRspecLAB). The precompiled version of MRspecLAB, along with the user manual and example datasets, the pre-defined pipelines (.pipe file) of the applications described in this manuscript, and the LCModel required control files (.control) and basis sets (.basis) are available on the zenodo repository (https://zenodo.org/records/14866163).

## 8 Author Contributions

Ying Xiao: Conceptualization, Data curation, Formal Analysis, Investigation, Methodology, Project administration, Software, Validation, Visualization, Writing – original draft, Writing – review & editing. Antonia Kaiser: Conceptualization, Data curation, Formal Analysis, Investigation, Methodology, Project administration, Resources, Supervision, Validation, Visualization, Writing – original draft, Writing – review & editing. Matthias Kockisch: Software, Writing – original draft. Alex Back: Investigation, Software, Validation, Writing – original draft. Robin Carlet: Software, Writing – original draft. Xinyu Liu: Resources, Validation, Writing – original draft. Zhiwei Huang: Resources, Validation, Writing – original draft. André Döring: Resources, Validation, Writing – original draft. Mark Widmaier: Resources, Validation, Writing – original draft. Lijing Xin: Conceptualization, Funding acquisition, Project administration, Supervision, Validation, Writing – original draft.

## 10 Acknowledgements

We acknowledge the MRI Platform of the FCBG (Fondation Campus Biotech Geneva) and the CIBM Center for Biomedical Imaging for providing expertise and resources to conduct this study. We acknowledge Dr. Yan Li (UCSF) and Dr. Anouk Schrantee (Amsterdam UMC) for providing the testing datasets. We also acknowledge the support from the Swiss National Science Foundation (grant no. 320030_189064 and 213769).

