## supplemental materials for "A graphical pipeline platform for MRS data processing and analysis: MRspecLAB"

### *Supplementary Material*

#### Supplement 1

A series of screenshots illustrating each processing step for another short-TE $^{1}$H dataset, which focuses on processing and quantifying short-TE $^{1}$H dataset, which acquired from the human brain on Siemens Terra.X 7T scanner with 1TX / 32RX Head Coil (Nova Medical). The example dataset were both in Siemens raw data format (.dat), included water-suppressed data acquired using the STEAM sequence (TE/TR = 4.5/4000 ms, bandwidth = 4 kHz, voxel size = $30 \times 30 \times 30$ mm$^3$, 32 averages) and unsuppressed water data obtained using the same sequence and parameters, from the human parietal lobe.

The screenshots demonstrate how MRspecLAB performs essential processing steps with the intuitive interface and the default-setting pipeline, such as (1) adaptive coil combination, (2) frequency and phase alignment, (3) eddy current correction, (4) bad average removal, (5) quality matrix, (6) spectral averaging, and (7) spectral fitting with LCModel. The results, including figures of processed data, fitting diagnostics, and quantification outputs, were generated and stored automatically.

(1) Adaptive coil combination: The input raw data for this step consists of uncombined signals from 32 individual receiver channels. During the adaptive coil combination process, MRspecLAB automatically combines these signals based on the input unsuppressed water data. The algorithm takes into account variations in coil sensitivities and phases across channels, applying phase correction and weighting to achieve the optimal combination. In the central plot panel, the data visualization of this step is organized into two rows. The first row displays the time-domain data, allowing users to observe the raw signal and its changes throughout this step. The second row showcases the frequency-domain data, presenting the spectra before and after variation corrections, which provides a clear, side-by-side comparison of the data before and after combination.

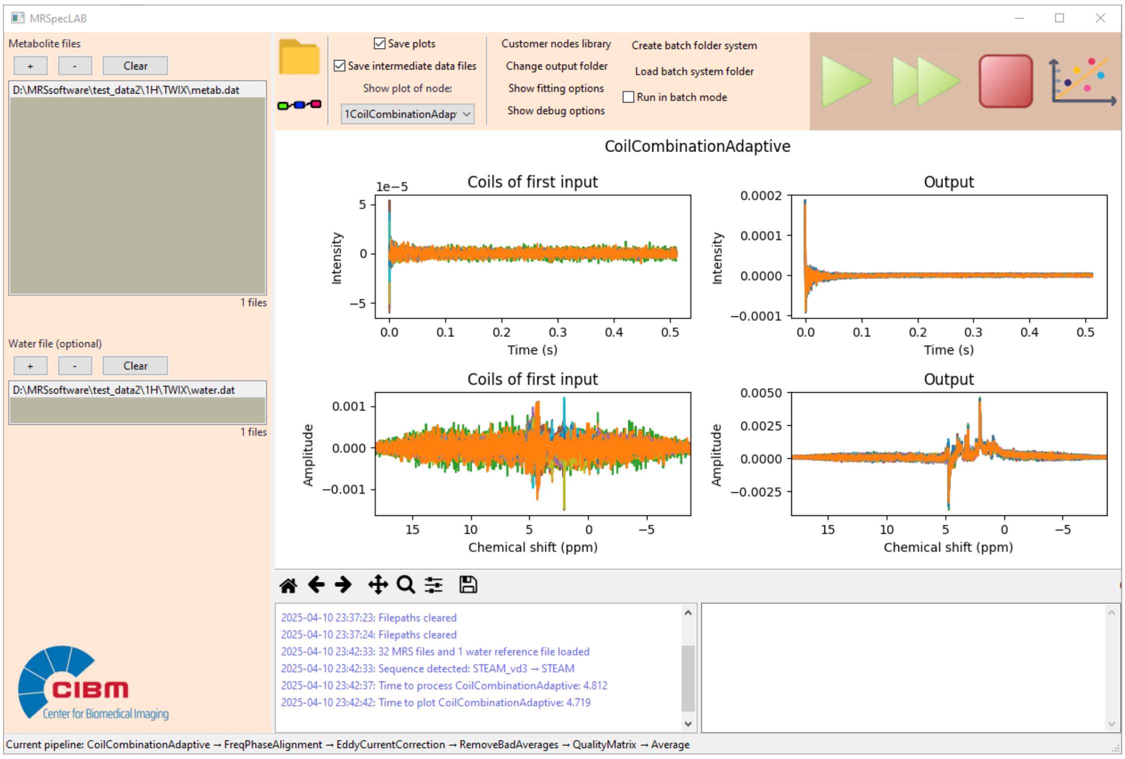


(2) Frequency and phase alignment: The data after coil combination proceeds to the alignment of individual transients utilizing the N-acetylaspartate (NAA) peak at 2.02 ppm as the reference. Three-fold zero padding was applied to the input data, followed by broadening using a Lorentzian function with a linewidth of 5 Hz. These preprocessing steps, illustrated in the left figure of the second row, were performed to enable accurate estimation of frequency and phase drifts in the dataset.

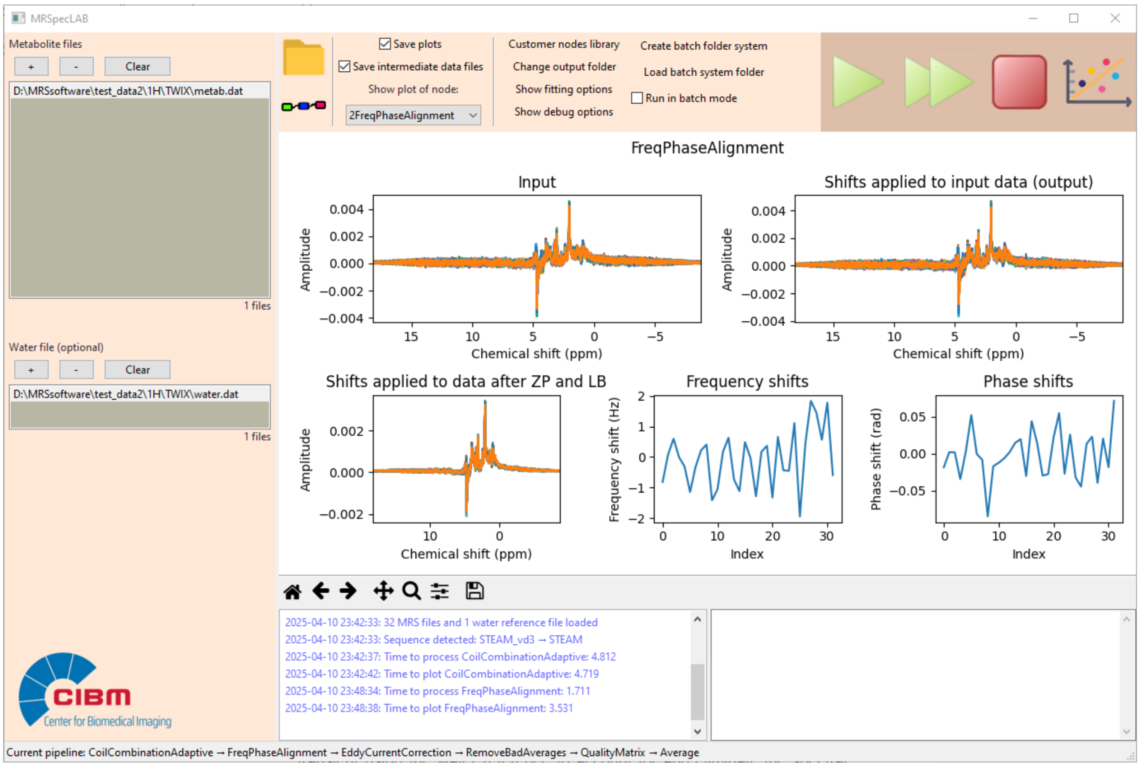

(3) Eddy current correction: Eddy current correction was performed on each transient using the water reference to account for and eliminate the spectral shape distortions. This step ensures improved spectral accuracy by correcting distortions caused by eddy currents. The non-linear phase before correction and the corrected phase after applying the correction are displayed in the figures.

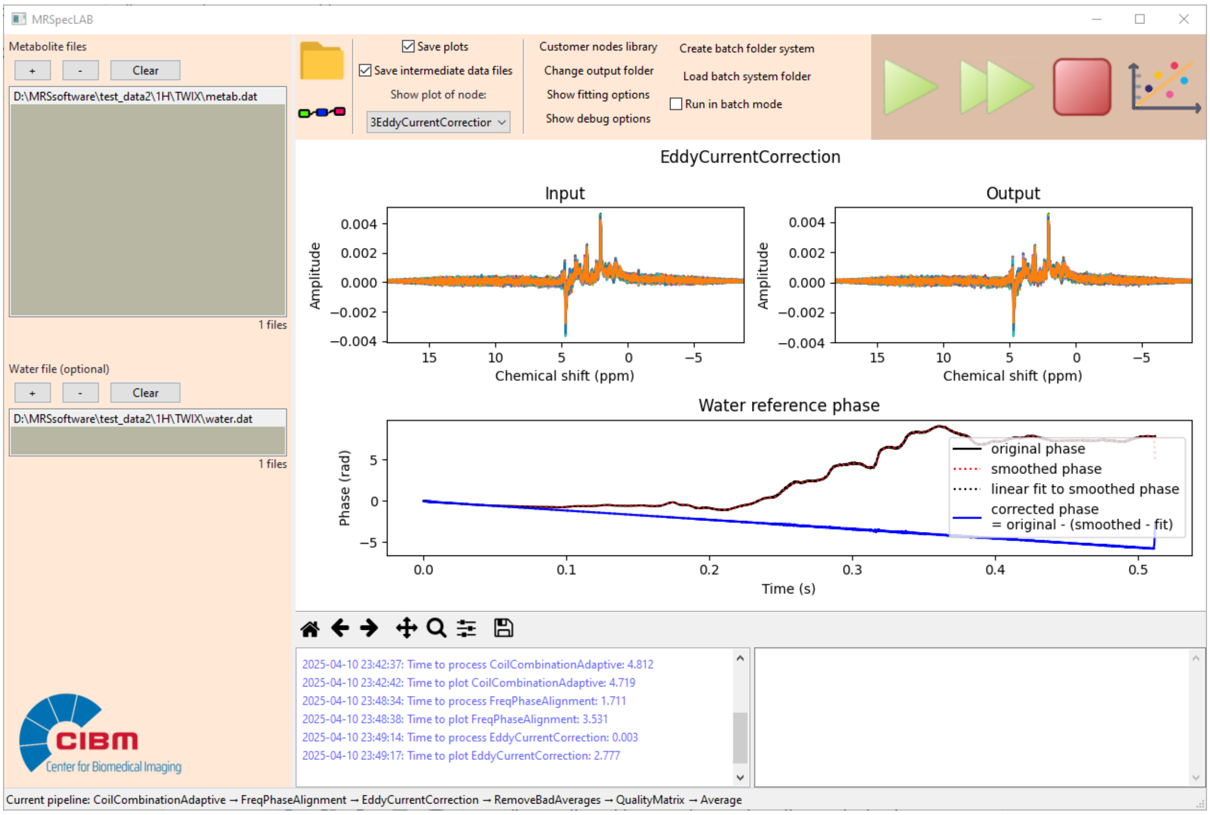


(4) Bad average removal: Following the eddy current correction, the similarity of each transient is evaluated to detect any deviations from the expected spectral pattern. Outlier transients, which may arise from factors such as hardware or motion, are identified and removed if they do not meet the similarity criteria, ensuring the quality of the dataset. In this particular dataset, no bad averages (shown in red, if any) were detected, so all transients were deemed acceptable (shown in gray) and moved forward to the next step in the processing pipeline.

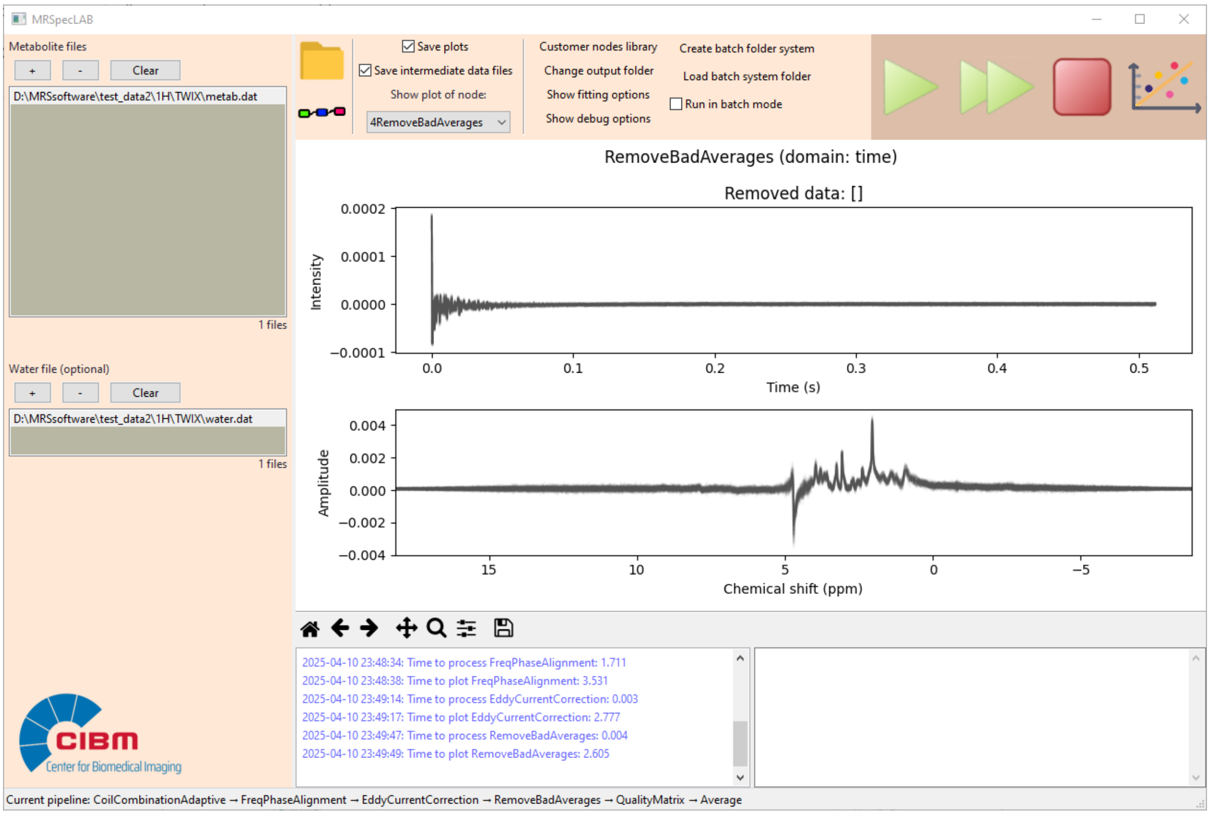


(5) Quality matrix: assess the quality of processed spectral data by calculating two key metrics: the SNR across the individual transients and the water linewidth. The SNR is computed for each spectrum to evaluate the signal strength of the NAA peak at 2.02 ppm relative to the noise from 0 to 0.5 ppm, indicating data quality. The water linewidth is estimated by fitting a Gaussian function to the water reference spectrum, allowing the calculation of the Full Width at Half Maximum (FWHM).

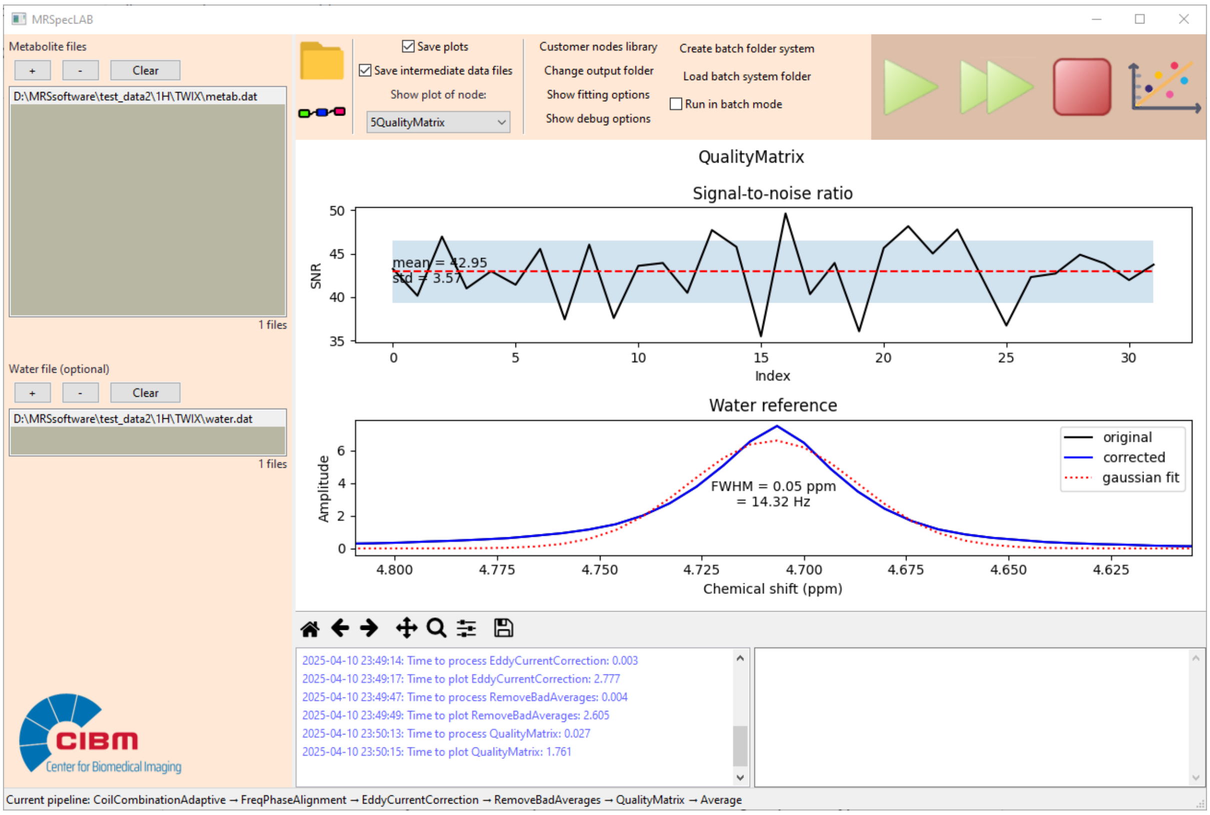


(6) Spectral averaging: The remaining good transients are averaged together. The left column of the figure shows the time-domain data, while the right column displays the spectra before and after averaging.

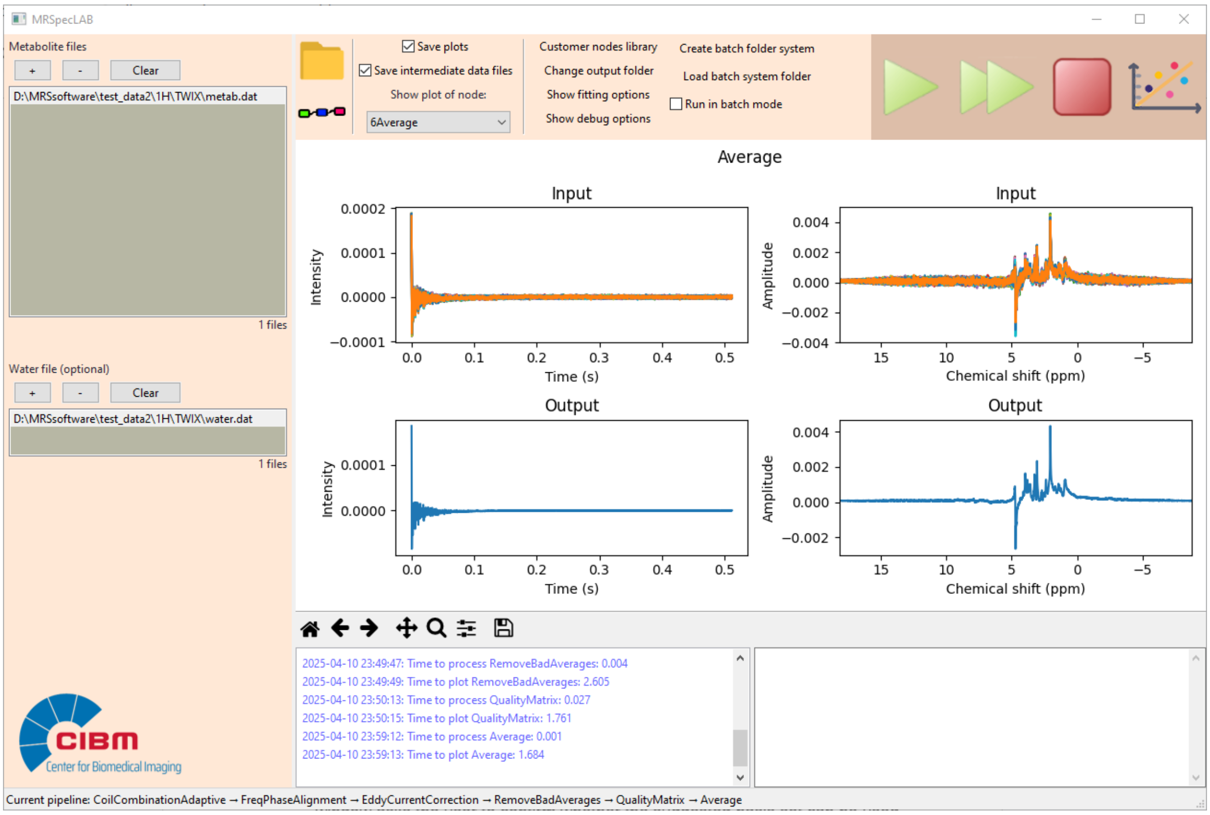

(7) Spectral fitting: If no specific basis set is selected by the user, MRspecLAB automatically detects the sequence information, including sequence type and scanning parameters such as TE, and attempts to match the data with a suitable basis set from the default library. Once a basis set is found, a prompt window asks the user to confirm whether the suggested basis set can be used for data fitting. Together with the fitting parameters (specified in the LCModel .control file), the software initiates the LCModel fitting process for the processed data. The final individual spectrum is displayed in the central figure panel, allowing users to visually inspect the fitting results. The quantitative analysis results extracted from the COORD file are summarized in the right information panel, providing users with quick access to key outcomes. Additionally, all fitting results generated by LCModel are systematically organized in the output folder, enabling users to review and utilize the results later as needed.

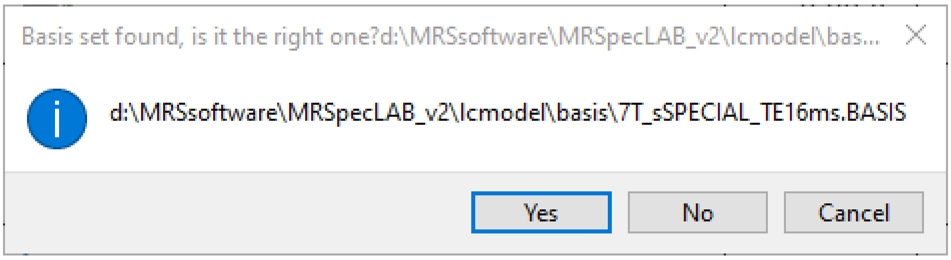


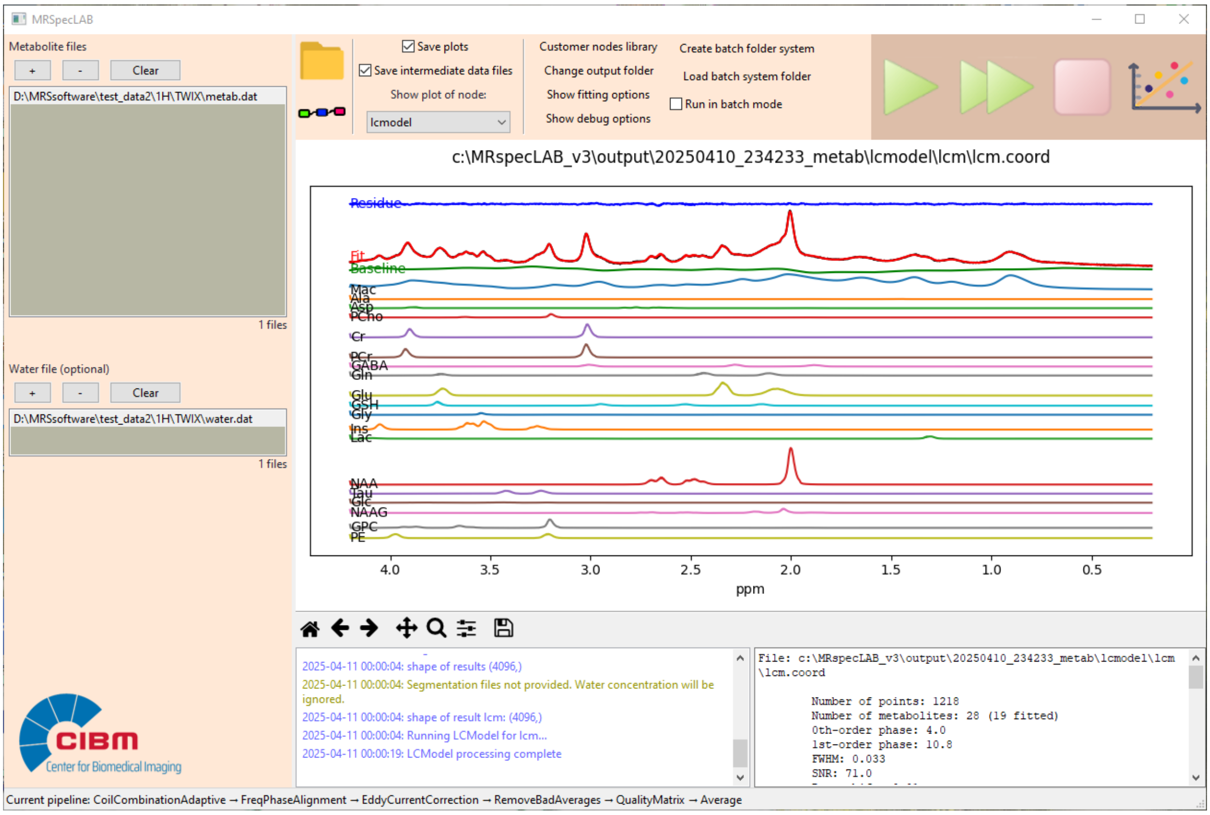

**Supplement 2**

MRspecLAB includes a default .control template for short-TE $^1$H MRS data quantification. Key parameters such as DELTAT, HZPPPM, and NUNFIL, which depend on the sequence and acquisition settings, are automatically extracted from the input data header, so users don't need to manually adjust these values when analyzing different datasets. Other fitting parameters are predefined in the default .control template and used during the fitting process. If users wish to fit with different parameter configurations, they can create and upload their own .control template through the fitting options before running the analysis. MRspecLAB will then automatically generate new .control files based on the uploaded template for each individual dataset. Additionally, users do not need to specify the directory or name of the input data in this file, as MRspecLAB directly links to the processed data, which allows to simplify the workflow and ensure compatibility across various datasets.

```
$LCMODL

 KEY= 210387309

 OWNER= 'MetMRS, CIBM, EPFL'

 DELTAT= 0.000125

 HZPPPM= 297.2041

 NUNFIL= 4096

 PPMEND= 0.2

 PPMST= 4.2

 NEACH= 999

 LPS= 8

 NSIMUL= 0

 ECHOT= 4.5

 DKNTMN= 0.2

 PPMSHF= 0

 NUSE1= 4

 CHUSE1(1)= 'NAA'

 CHUSE1(2)= 'Cr'

 CHUSE1(3)= 'Glu'
```

CHUSE1(4)= 'Ins'

NCOMBI= 4

CHCOMB(1)= 'Glu+Gln'

CHCOMB(2)= 'Cr+PCr'

CHCOMB(3)= 'NAA+NAAG'

CHCOMB(4)= 'GPC+PCho'

VITRO = F

ATTH2O= 1

WCONC= 44444

DEGZER= 0

DOECC = F

DOWS = T

DOREFS = T

SHIFMN = -0.2,-0.1

SHIFMX = 0.3,0.3

$END

**Supplement 3**

fMRS dataset processing pipeline and results

1) Coil combination

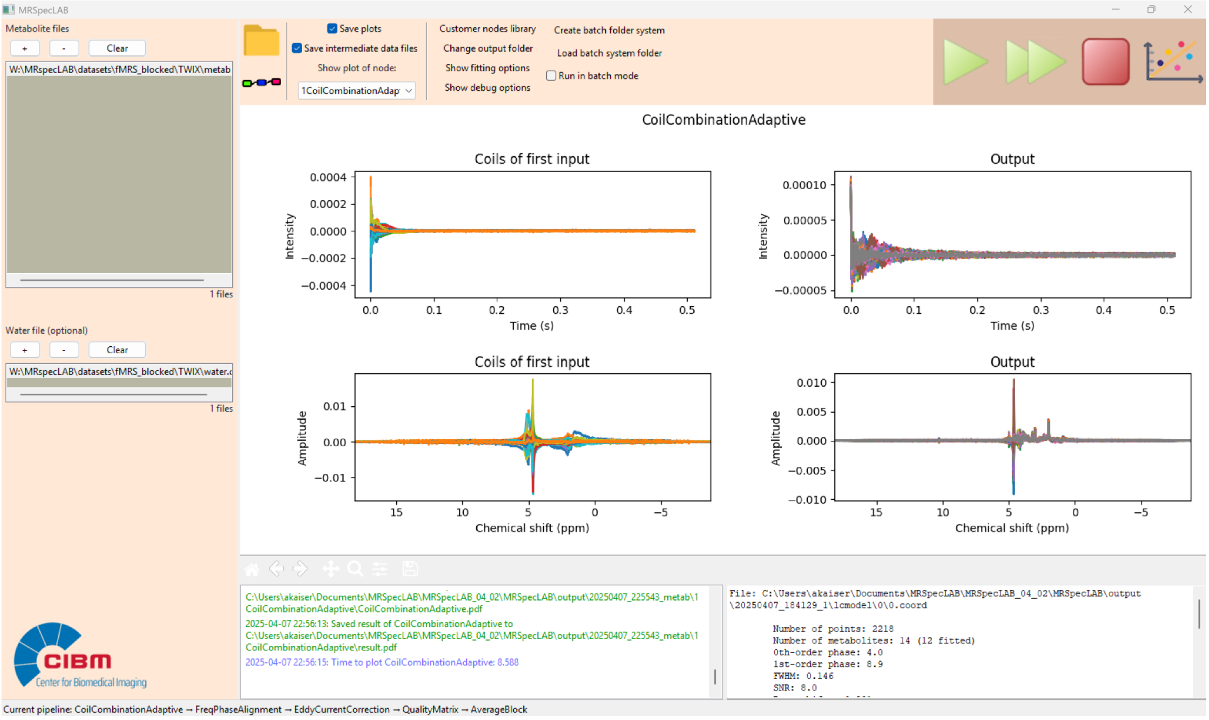


2) Frequency and phase alignment

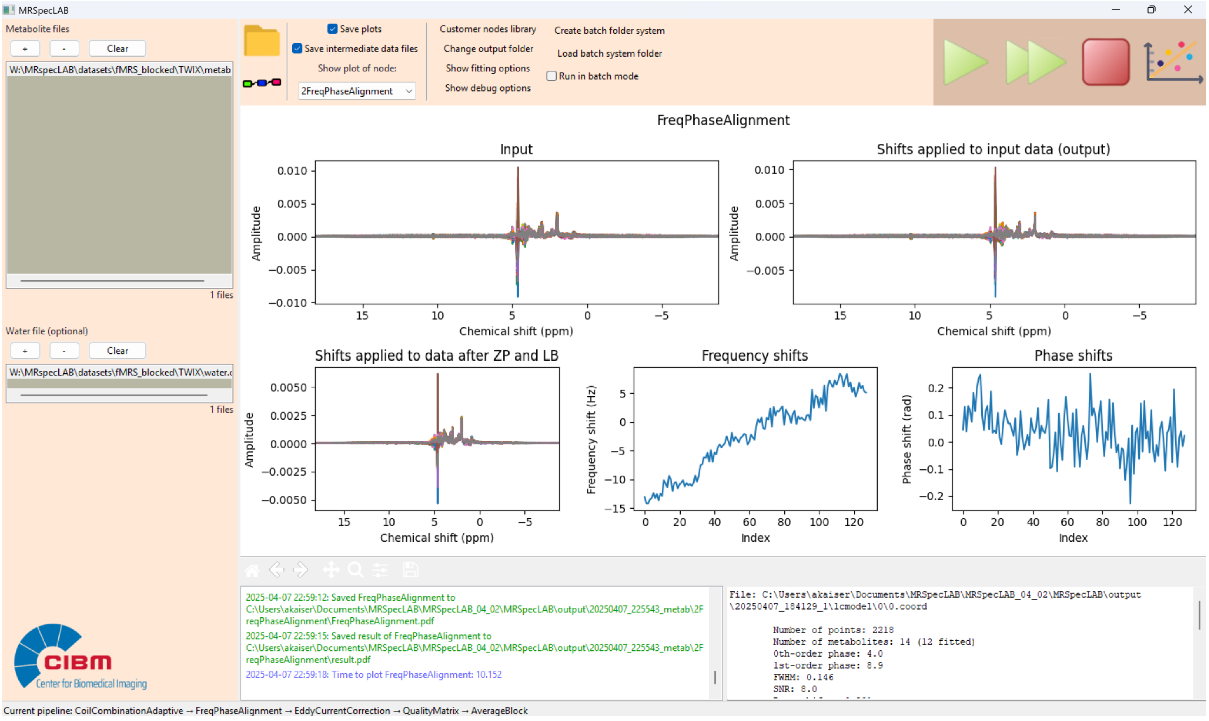


3) Eddy current correction

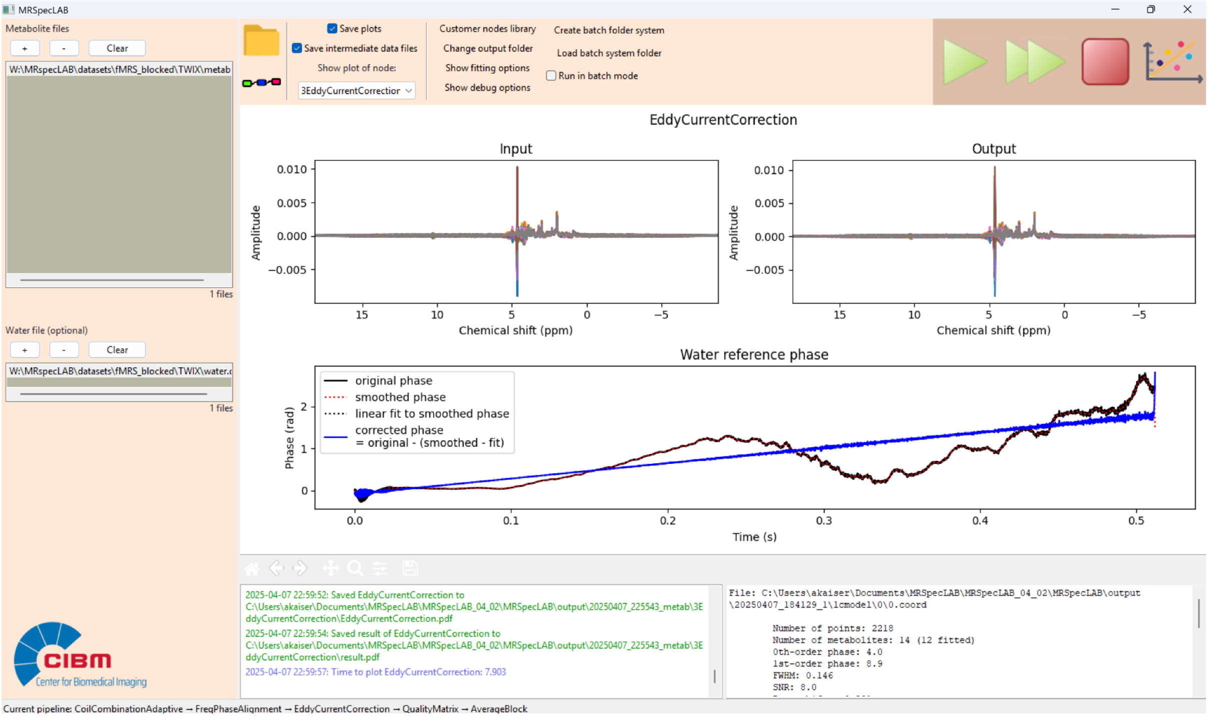


4) Quality matrix outcome

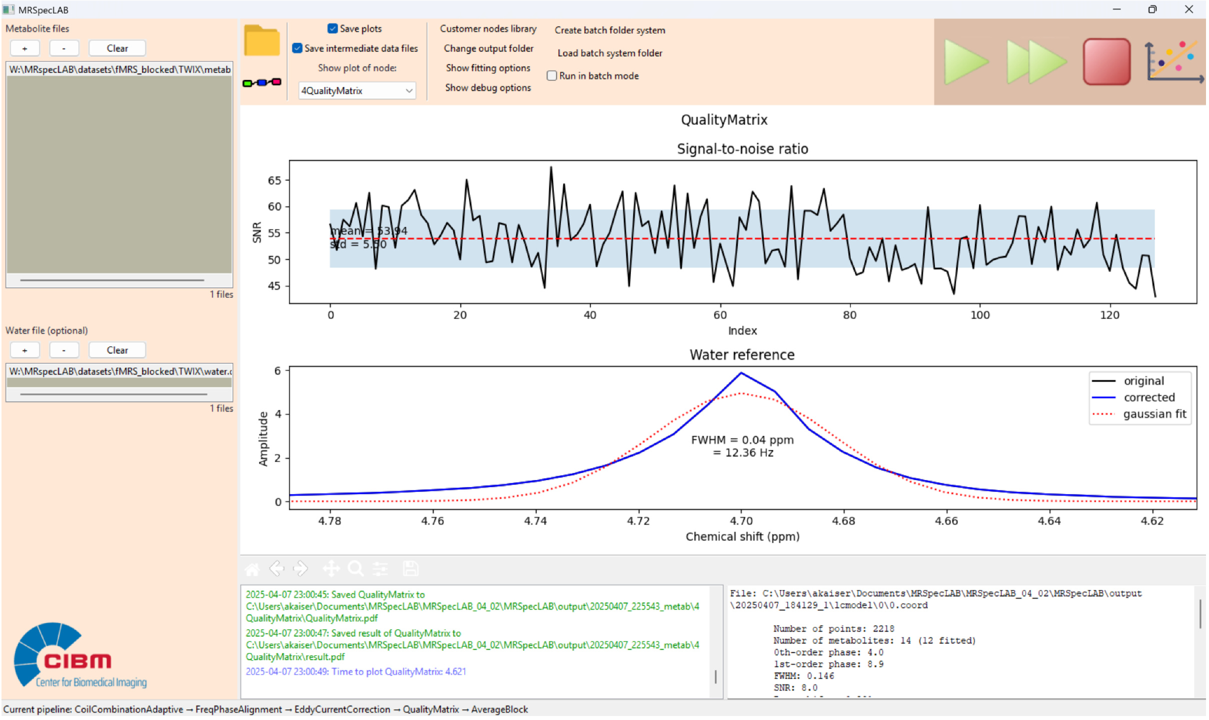


5) Block averaging

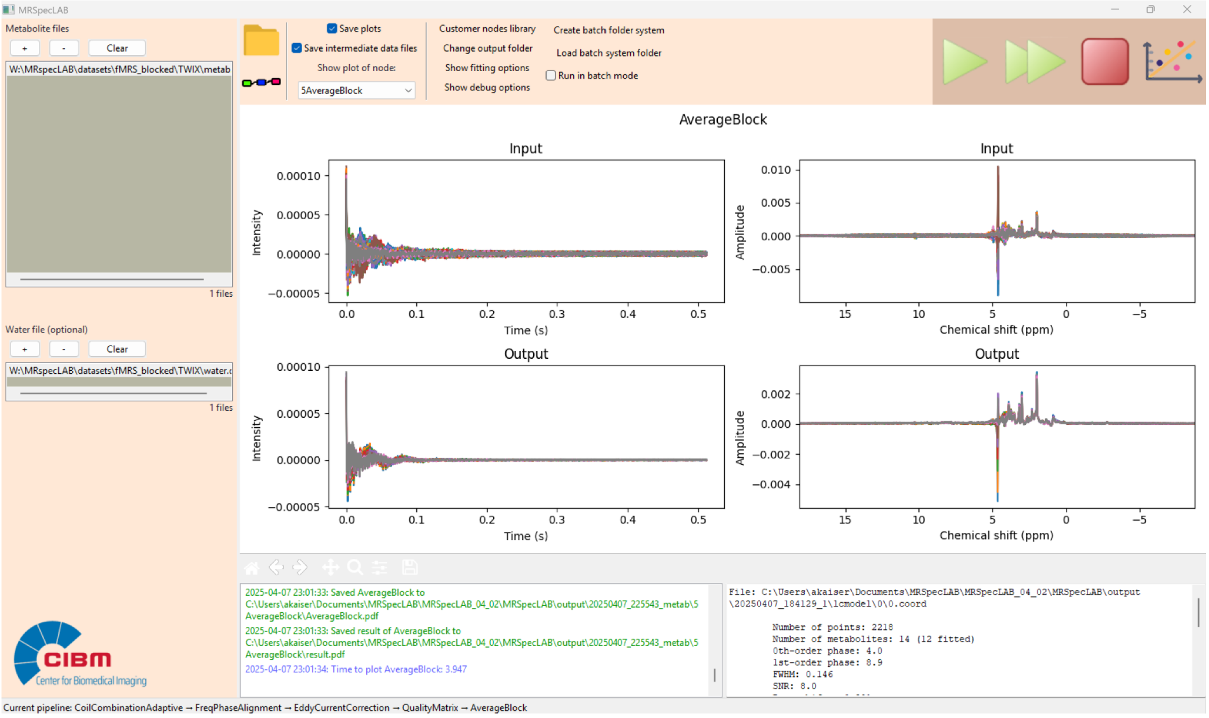


6) LCModel fitting per average

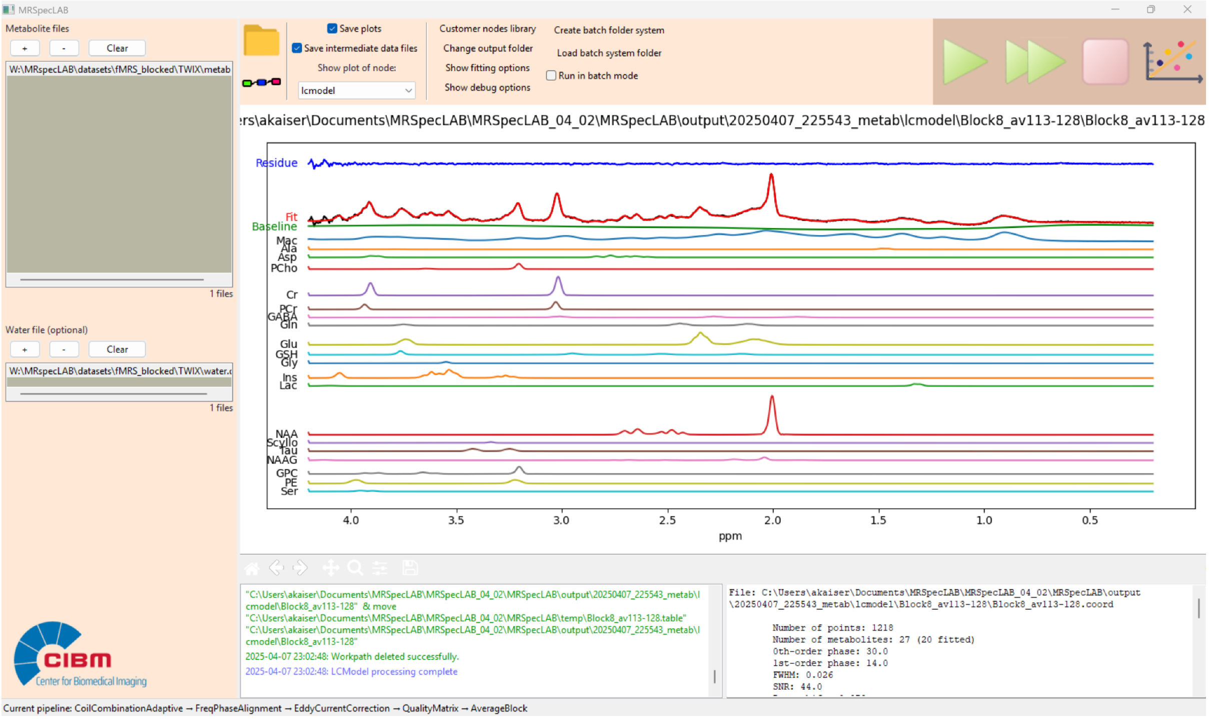


#### Supplement 4

Comparison of post-processed spectra (n=5) using MRspecLAB and FID-A.

The input data acquired with and without water suppression in TWIX format (sequence: sSPECIAL, VOI = 25 × 30 × 30 $cm^3$, TE/TR = 16/4000 ms, BW = 4 kHz, NT = 16 for water-suppressed data, and NT = 2 for unsuppressed water data).

This analysis provides a comparison of the above mentioned 1H MRS spectra processed with MRspecLAB and FID-A using identical raw data acquired in Siemens TWIX format. Both analysis pipelines included the same preprocessing steps: coil combination based on the water reference signal, frequency and phase alignment, eddy current correction, outlier average removal, and signal averaging.

For FID-A, the processing pipeline included the following standard functions: io_loadspec_twix → op_addrcvrs → op_alignAverages → op_rmbadaverages → op_averaging → op_ecc.

For MRspecLAB, the corresponding nodes included were: CoilCombinationAdaptive → FreqPhaseAlignment → EddyCurrentCorrection → RemoveBadAverages → Quality Matrix → Average.

Visual inspection of the processed spectra shows high consistency in spectral features across both methods, with strong similarity in peak shapes, baseline stability, and noise characteristics (Figure below). With participants depicted per row (P1-5) and colors indicating the toolbox used for preprocessing (pink: FID-A, blue: MRspecLAB).

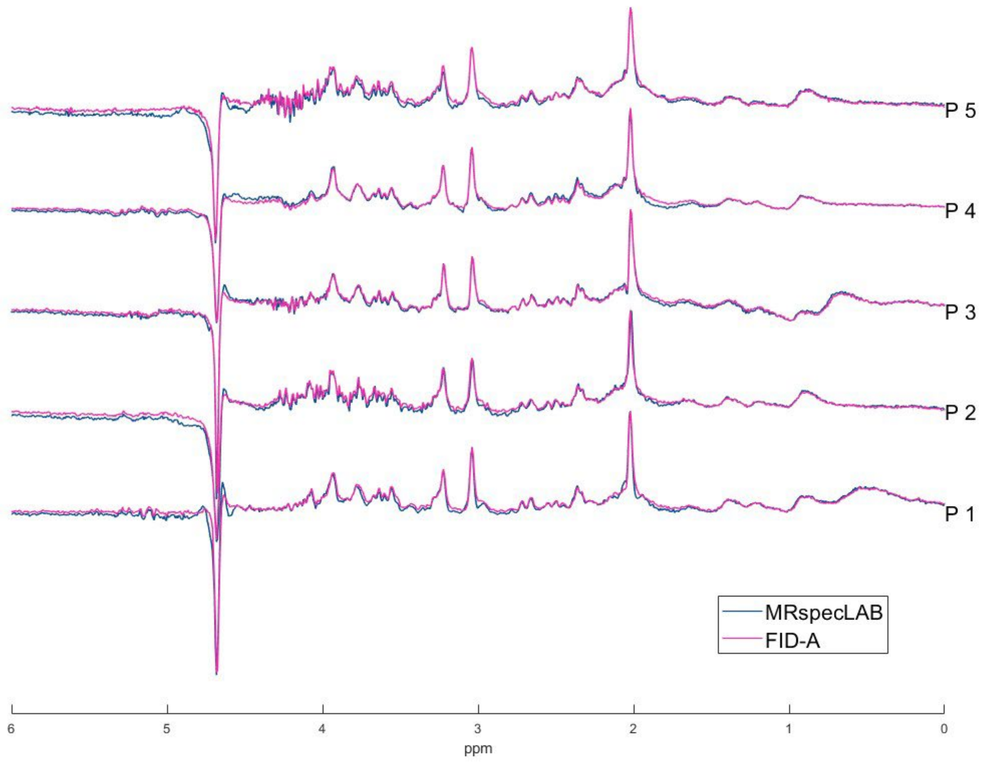


Spectral quantification was performed using LCModel version 6.3 with the same basis set and control file for both toolboxes. Major metabolite concentrations—such as NAA, Cr, and Glu—

are clearly and consistently represented in the fitting results from both tools (no tissue correction).

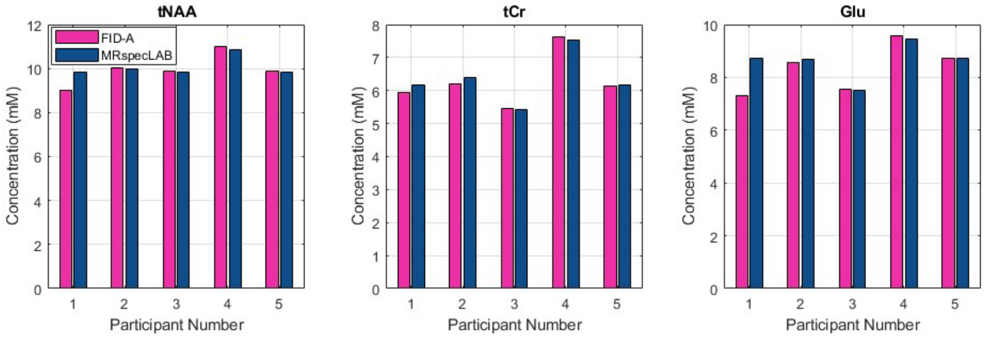


#### Supplement 5

$^{31}$P dataset processing and results

1) Apodization of 5 Hz

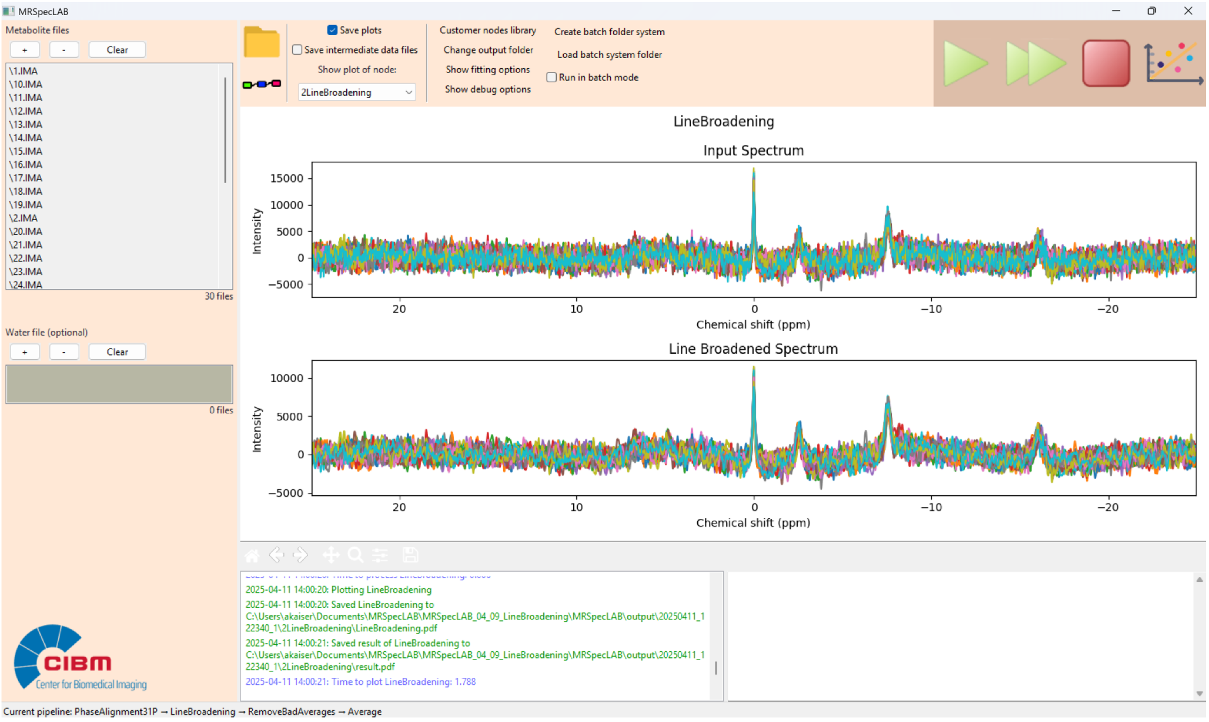


2) Averaging

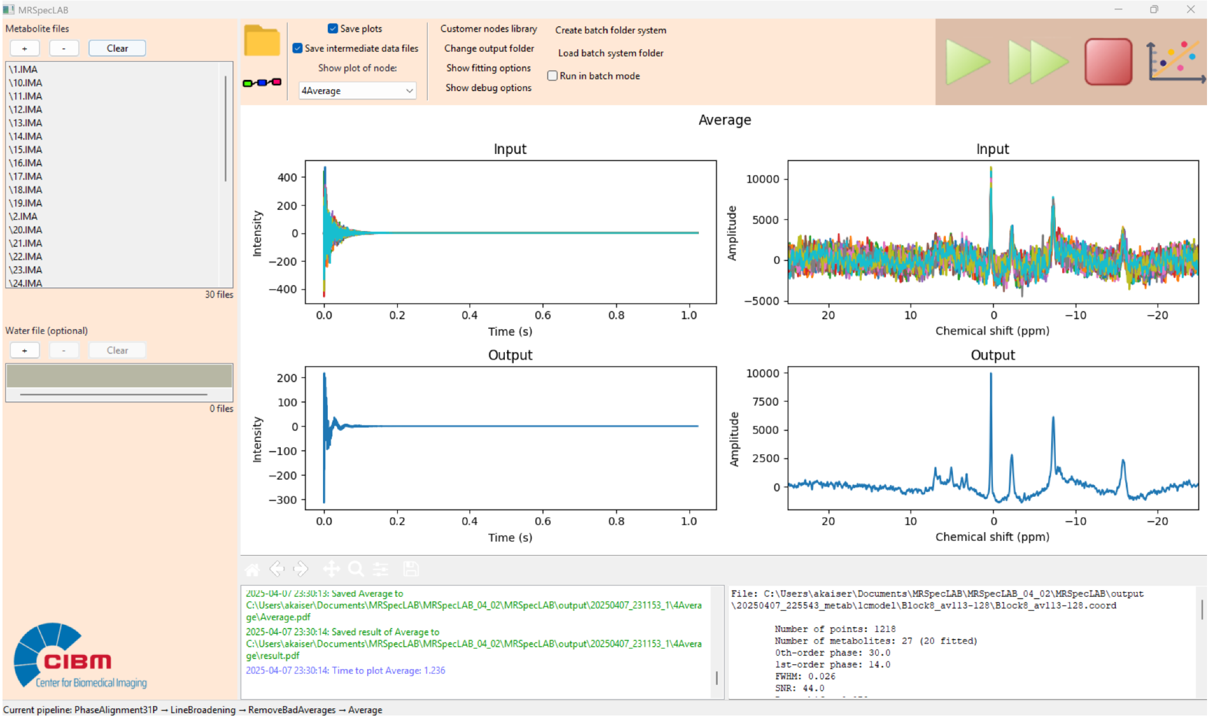


#### 3) LCModel fitting

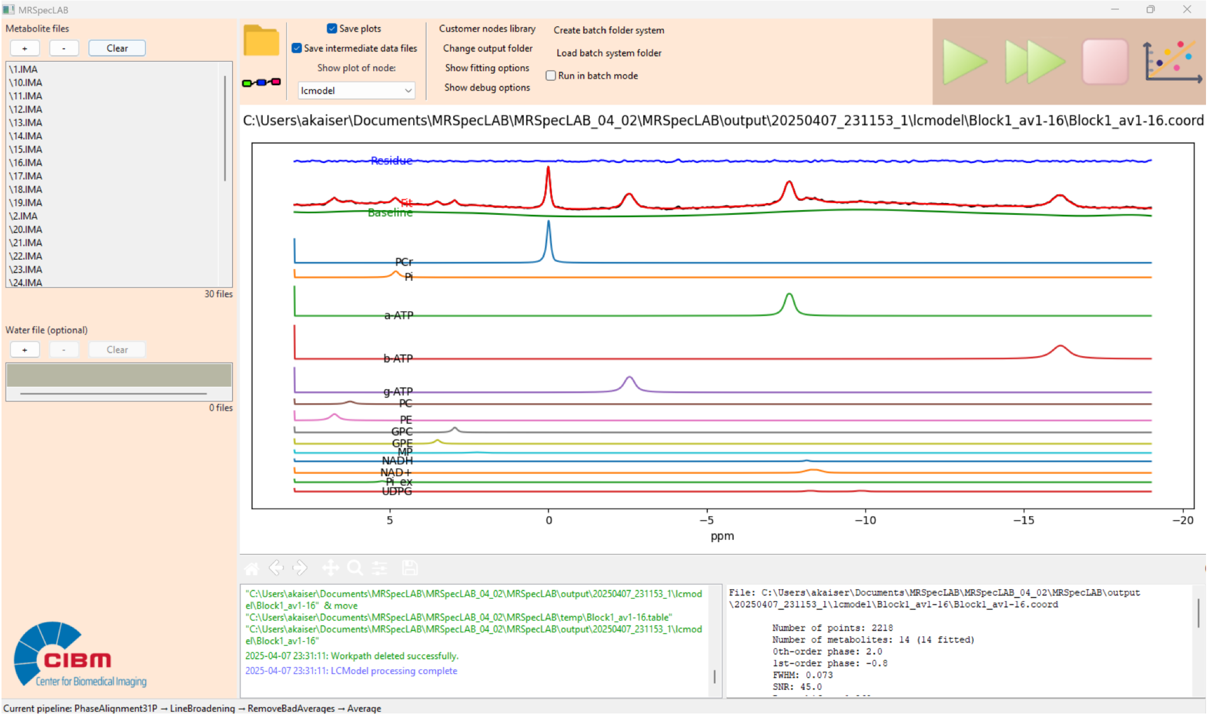


#### Supplements 6

$^{31}$P CSI data processing and fitting

1) Hanning filter

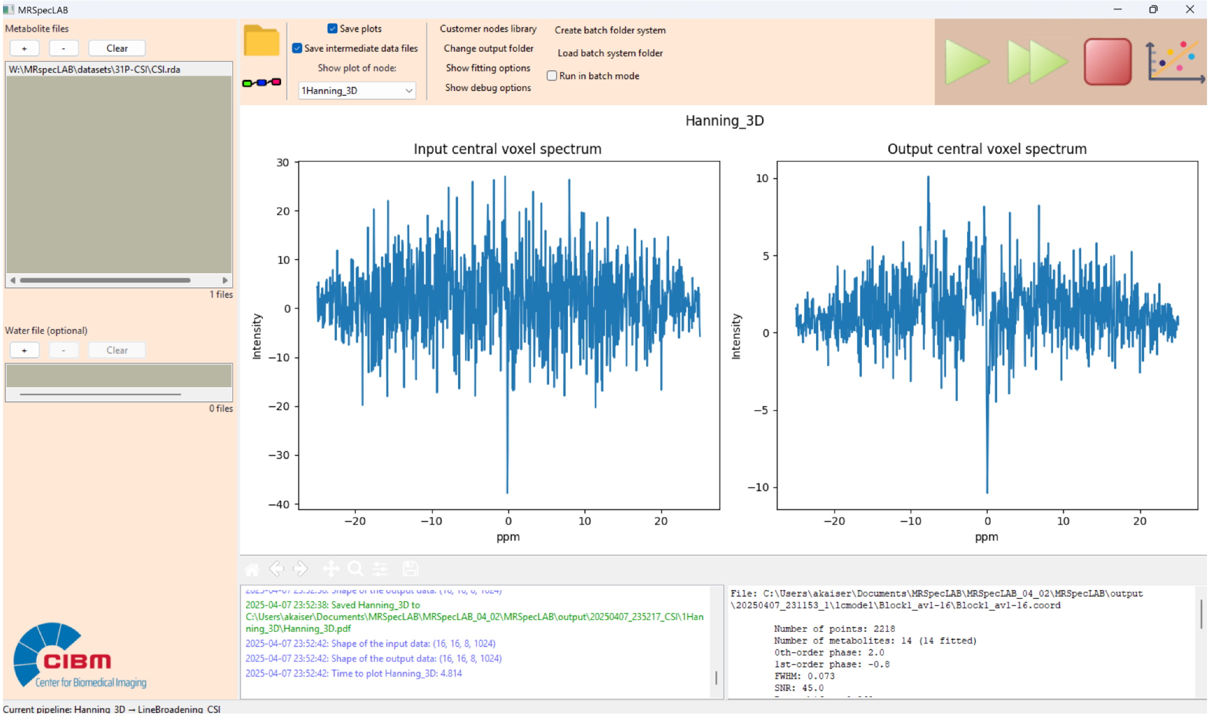


2) Apodization 5 Hz

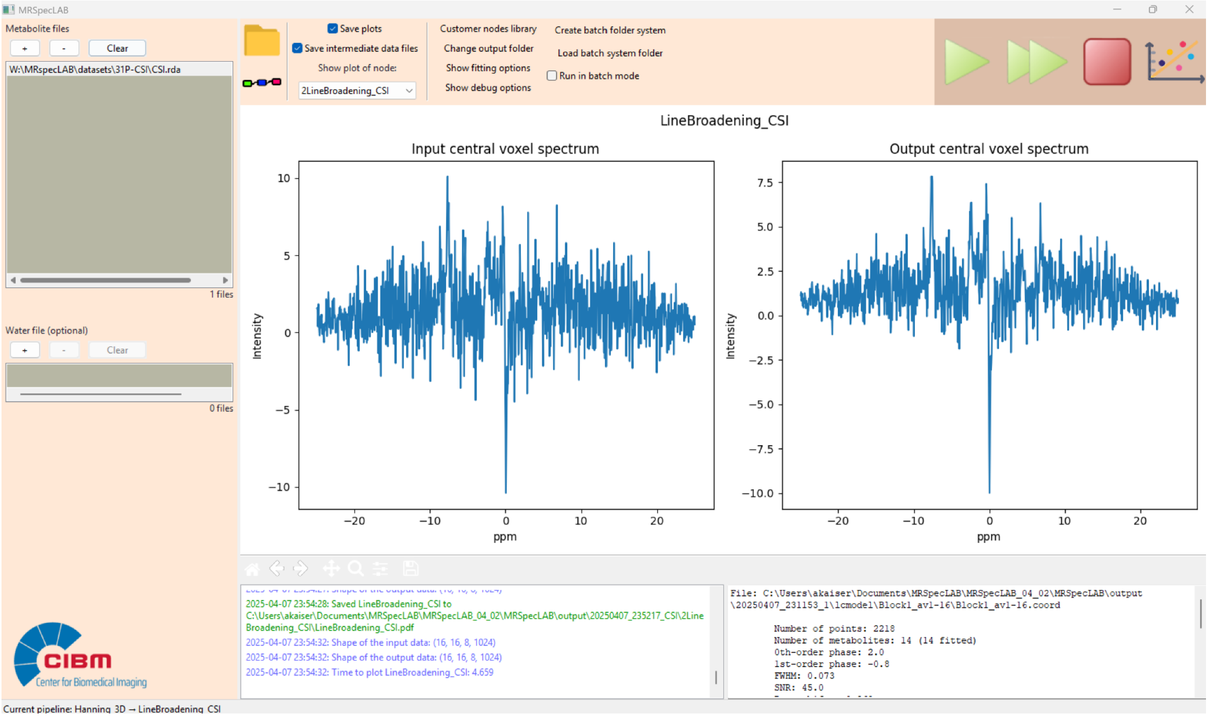


3) LCModel fitting (voxel by voxel)

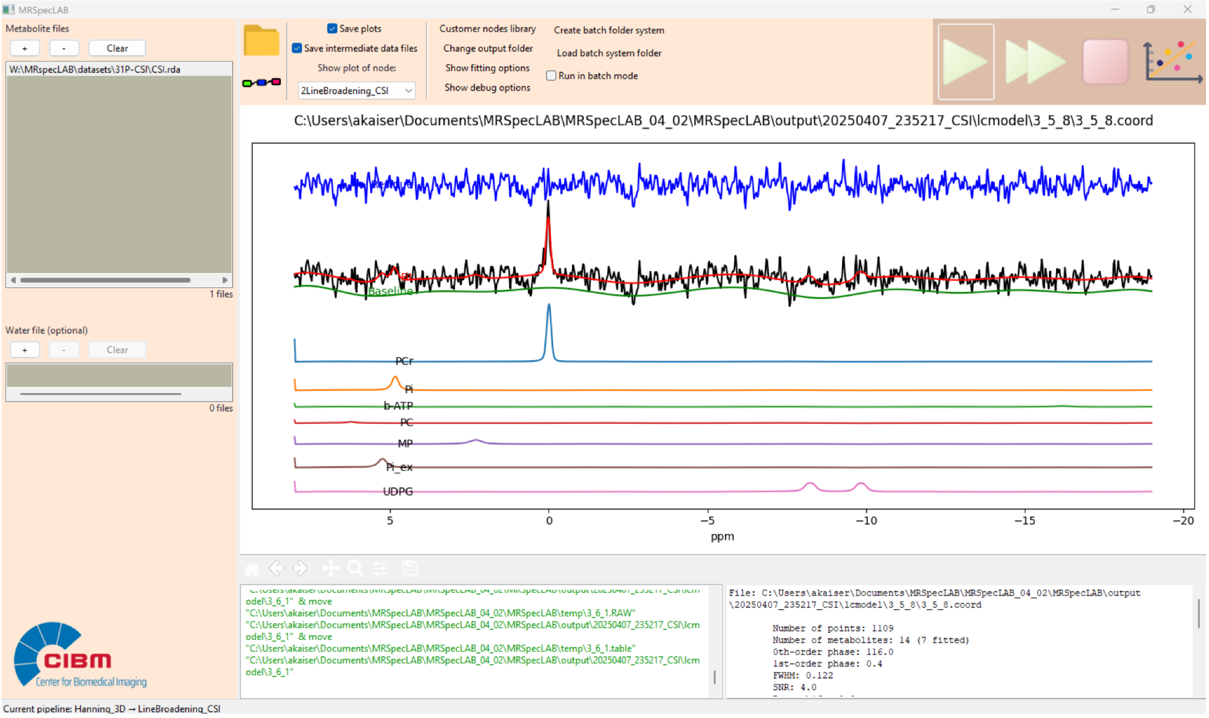


**Supplement 7**

Below is an example code for creating a custom node that applies Gaussian line broadening:

```
# Import necessary APIs and libraries

import processing.api as api

import numpy as np


# Define the custom node class

class LineBroadening_Gaussian(api.ProcessingNode):

    def __init__(self, nodegraph, id):

        # Meta information about the node

        self.meta_info = {

            "label": "Line Broadening (Gaussian)",

            "author": "MetMRS",

            "description": "Applies Gaussian line broadening to spectral data",
```

```
    }
    # Define adjustable parameters
    self.parameters = [
        api.IntegerProp(
            idname="Gaussian_lw_hz",
            default=5,  # Default linewidth (Hz)
            min_val=1,  # Minimum value
            max_val=50,  # Maximum value
            fpb_label="Linewidth (Hz)"  # Label for the GUI
        )
    ]
    super().__init__(nodegraph, id)  # Initialize the parent class
    self.plotSpectrum = False  # Disable spectrum plotting by default

def process(self, data):
    # Create Gaussian apodization function
    self.exp = np.exp(-((data["input"][0].time_axis() * np.pi *
                        self.get_parameter("Gaussian_lw_hz")) /
                        (2 * np.sqrt(np.log(2)))) ** 2)
    output = []
    self.dmax = 0
    for d in data["input"]:
        # Apply the defined function
        output.append(d.inherit(d * self.exp))
        self.dmax = max(self.dmax, np.max(d))  # Track max value for plotting
    data["output"] = output
```

```
    def plot(self, figure, data):
        # Plot input, apodization function, and output
        figure.suptitle(self.__class__.__name__)
        ax = figure.add_subplot(2, 1, 1)
        for d in data["input"]:
            ax.plot(d.time_axis(), np.real(d))
        ax.plot(d.time_axis(), self.exp * self.dmax, ':k')
        ax.set_xlabel('Time (s)')
        ax.set_ylabel('Signal Intensity (a.u.)')
        ax.set_title("Input and Apodization Function")
        ax = figure.add_subplot(2, 1, 2)
        for d in data["output"]:
            ax.plot(d.time_axis(), np.real(d))
        ax.set_xlabel('Time (s)')
        ax.set_ylabel('Signal Intensity (a.u.)')
        ax.set_title("Output")
        figure.tight_layout()

# Register the custom node to make it available in the node library
api.RegisterNode(LineBroadening_Gaussian, "LineBroadening_Gaussian")
```

**Supplements 8**

Output is organized by numbered folders per processing step. Each folder contains a diagnostic plot for quality control of the respective step, and a results figure showing the resulting spectra (.PDF). Additionally, if selected, the results of every processing step are saved in a ‘’data’’ folder as LCModel .RAW output and NIFTI-MRS. The LCModel output folder contains several files: .control: Contains all setting parameters for the fit; .coord: Includes fitted spectral curves, quality control parameters, and final fit results; .csv: Summarizes quantification results per metabolite; .ps: Provides a visual representation of all fitted spectra; .print: Contains detailed analysis information, including metabolite concentrations and correlations; .nii: Stores fitted metabolite maps in NIfTI format; .H2O: Contains raw water spectral data; .table: Presents quantification results in tabulated format; .RAW: Stores raw metabolite spectral data in the time domain (Provencher SW. NMR Biomed. 2001;14(4):260–4.).

```
OUTPUT_FOLDER/
├── pipeline.pipe              # Saves the pipeline settings used for processing
├── MRSinMRS_table.csv         # Standardized table for MRS acquisition and processing parameters
├── 1CoilCombination/          # First processing step (Coil Combination)
│   ├── data/
│   │   ├── <FILENAME>.RAW     # Raw metabolite spectral data (ASCII format, if selected)
│   │   ├── <FILENAME>.nii     # Intermediate results in NIfTI-MRS format (if selected)
│   ├── results.pdf            # Results of Coil Combination processing
│   ├── CoilCombination.pdf    # Quality control output for Coil Combination
├── 2Preprocessing/            # Second processing step (Preprocessing)
│   ├── data/
│   │   ├── <FILENAME>.RAW     # Processed spectral data (if selected)
│   │   ├── <FILENAME>.nii     # Processed spectral data in NIfTI format (if selected)
│   ├── results.PDF            # Results of Preprocessing
│   ├── Preprocessing.pdf      # Quality control output for Preprocessing
├── 3Averaging/                # Third processing step (Averaging)
│   ├── data/
│   │   ├── <FILENAME>.RAW     # Averaged spectral data (if selected)
│   │   ├── <FILENAME>.nii     # Averaged spectral data in NIfTI format (if selected)
│   ├── results.PDF            # Results of Averaging
│   ├── Averaging.pdf          # Quality control output for Averaging
├── LCModel/                   # LCModel output folder (at the same level as processing steps)
│   ├── <FILENAME>.control     # LCModel control file with all fit settings
│   ├── <FILENAME>.basis       # Basis set used for spectral fitting
│   ├── <FILENAME>.coord       # Fitted spectral curves and quality control parameters
│   ├── <FILENAME>.csv         # Quantification results per metabolite
│   ├── <FILENAME>.ps          # Visual representation of all fitted spectra
│   ├── <FILENAME>.print       #Detailed analysis output including metabolite concentrations
│   ├── <FILENAME>.nii         # Fitted metabolite maps in NIfTI format
│   ├── <FILENAME>.table       # Quantification results in tabulated format
│   ├── <FILENAME>.H2O         # Raw water spectral data for quantification (optional)
│   ├── <FILENAME>.RAW         # Raw metabolite spectral data in the time domain
```

**Example output folder in Application 1:**

The data and figures from each step are organized in subfolders named after the corresponding processing nodes. A summary of the data information is provided in the MRSinMRS table. Additionally, the processing pipeline is stored within the output folder.

| Name | Date modified | Type | Size |
|---|---|---|---|
| 1CoilCombinationAdaptive | 10/04/2025 23:42 | File folder | |
| 2FreqPhaseAlignment | 10/04/2025 23:48 | File folder | |
| 3EddyCurrentCorrection | 10/04/2025 23:49 | File folder | |
| 4RemoveBadAverages | 10/04/2025 23:49 | File folder | |
| 5QualityMatrix | 10/04/2025 23:50 | File folder | |
| 6Average | 10/04/2025 23:59 | File folder | |
| lcmodel | 11/04/2025 00:00 | File folder | |
| MRSinMRS_table.csv | 10/04/2025 23:42 | Microsoft Excel C... | 4 KB |
| pipeline.pipe | 10/04/2025 23:59 | PIPE File | 1 KB |
| Result.pdf | 11/04/2025 00:00 | Adobe Acrobat D... | 72 KB |

For example, in 1CoilCombinationAdaptive, we will have:

| Name | Date modified | Type | Size |
|---|---|---|---|
| data | 10/04/2025 23:42 | File folder | |
| CoilCombinationAdaptive.pdf | 10/04/2025 23:42 | Adobe Acrobat D... | 3.532 KB |
| result.pdf | 10/04/2025 23:42 | Adobe Acrobat D... | 2.107 KB |

Once "Save plots" is clicked, all time and frequency domain figures will be saved in PDF format under each step's folder, named "results.pdf" for the user's review. Additionally, a PDF with the same name as the processing node will be saved, containing the figure currently displayed on the main canvas.

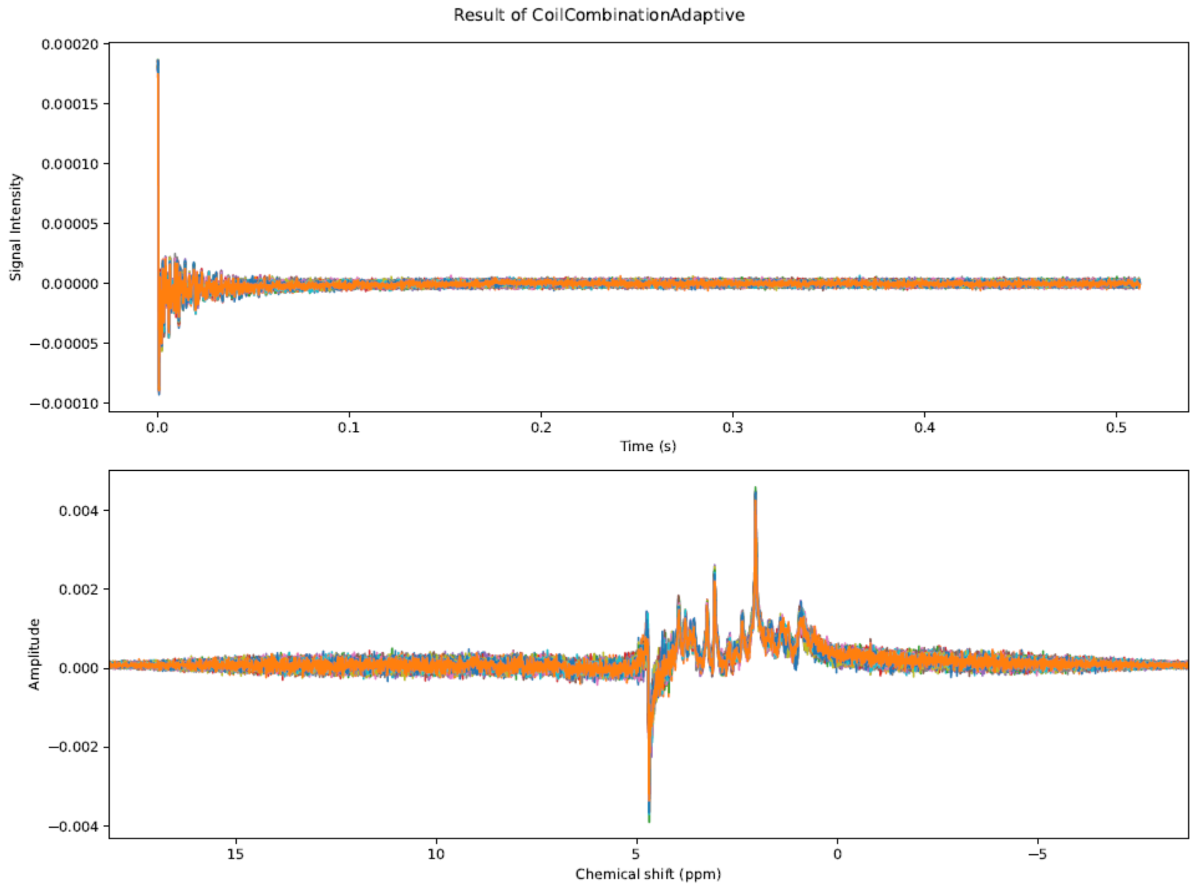


Once "Save intermediate datafiles" is clicked, all intermediate time domain data will be stored in .RAW and .nii formats. These files can be found in the subfolder named "data" within each processing step's folder.

| Name | Date modified | Type | Size |
|---|---|---|---|
| 0.nii | 10/04/2025 23:42 | NIfTI | 33 KB |
| 0.RAW | 10/04/2025 23:42 | RAW File | 129 KB |
| 1.nii | 10/04/2025 23:42 | NIfTI | 33 KB |
| 1.RAW | 10/04/2025 23:42 | RAW File | 129 KB |
| 2.nii | 10/04/2025 23:42 | NIfTI | 33 KB |
| 2.RAW | 10/04/2025 23:42 | RAW File | 129 KB |
| 3.nii | 10/04/2025 23:42 | NIfTI | 33 KB |
| 3.RAW | 10/04/2025 23:42 | RAW File | 129 KB |
| 4.nii | 10/04/2025 23:42 | NIfTI | 33 KB |
| 4.RAW | 10/04/2025 23:42 | RAW File | 129 KB |
| 5.nii | 10/04/2025 23:42 | NIfTI | 33 KB |
| 5.RAW | 10/04/2025 23:42 | RAW File | 129 KB |
| 6.nii | 10/04/2025 23:42 | NIfTI | 33 KB |
| 6.RAW | 10/04/2025 23:42 | RAW File | 129 KB |
| 7.nii | 10/04/2025 23:42 | NIfTI | 33 KB |
| 7.RAW | 10/04/2025 23:42 | RAW File | 129 KB |
| 8.nii | 10/04/2025 23:42 | NIfTI | 33 KB |
| 8.RAW | 10/04/2025 23:42 | RAW File | 129 KB |
| 9.nii | 10/04/2025 23:42 | NIfTI | 33 KB |
| 9.RAW | 10/04/2025 23:42 | RAW File | 129 KB |
| 10.nii | 10/04/2025 23:42 | NIfTI | 33 KB |
| 10.RAW | 10/04/2025 23:42 | RAW File | 129 KB |
| 11.nii | 10/04/2025 23:42 | NIfTI | 33 KB |
| 11.RAW | 10/04/2025 23:42 | RAW File | 129 KB |
| 12.nii | 10/04/2025 23:42 | NIfTI | 33 KB |
| 12.RAW | 10/04/2025 23:42 | RAW File | 129 KB |
| 13.nii | 10/04/2025 23:42 | NIfTI | 33 KB |
| 13.RAW | 10/04/2025 23:42 | RAW File | 129 KB |
| 14.nii | 10/04/2025 23:42 | NIfTI | 33 KB |
| 14.RAW | 10/04/2025 23:42 | RAW File | 129 KB |
| 15.nii | 10/04/2025 23:42 | NIfTI | 33 KB |
| 15.RAW | 10/04/2025 23:42 | RAW File | 129 KB |
| 16.nii | 10/04/2025 23:42 | NIfTI | 33 KB |
| 16.RAW | 10/04/2025 23:42 | RAW File | 129 KB |
| 17.nii | 10/04/2025 23:42 | NIfTI | 33 KB |
| 17.RAW | 10/04/2025 23:42 | RAW File | 129 KB |
| 18.nii | 10/04/2025 23:42 | NIfTI | 33 KB |
| 18.RAW | 10/04/2025 23:42 | RAW File | 129 KB |
